# Analysis of Collective Response Reveals that COVID-19-Related Activities Start From the End of 2019 in Mainland China

**DOI:** 10.1101/2020.10.14.20202531

**Authors:** Ji Liu, Tongtong Huang, Haoyi Xiong, Jizhou Huang, Jingbo Zhou, Haiyan Jiang, Guanhua Yang, Haifeng Wang, Dejing Dou

## Abstract

While the COVID-19 outbreak is making an impact at a global scale, the collective response to the pandemic becomes the key to analyzing past situations, evaluating current measures, and formulating future predictions. In this paper, we analyze the public reactions to the pandemic using search engine data and mobility data from Baidu Search and Baidu Maps respectively, where we particularly pay attentions to the early stage of pandemics and find early signals from the collective response to COVID-19. First, we correlate the number of confirmed cases per day to daily search queries of a large number of keywords through Dynamic Time Warping (DTW) and Detrended Cross-Correlation Analysis (DCCA), where the keywords top in the most critical days are believed the most relevant to the pandemic. We then categorize the ranking lists of keywords according to the specific regions of the search, such as Wuhan, Mainland China, the USA, and the whole world. Through the analysis on search, we succeed in identifying COVID-19 related collective response would not be earlier than the end of 2019 in Mainland China. Finally, we confirm this observation again using human mobility data, where we specifically compare the massive mobility traces, including the real-time population densities inside key hospitals and inter-city travels departing from/arriving in Wuhan, from 2018 to 2020. No significant changes have been witnessed before December, 2019.

## Introduction

As reported by the World Health Organization (WHO), a global pandemic, COVID-19, has rapidly spread all over the world since December, 2019, e.g., within China (*1*), the United States (*2*), European countries (*3*), etc. The COVID-19 pandemic is considered as the greatest challenge for humankind since World War II (*4*). The outbreak of COVID-19 has been closely monitored by governments, researchers, and digital tracing applications (*5, 6*). As early as January 2, 2020 research (*7*) has confirmed Huanan seafood market in Wuhan was epidemiologically associated with most (27 of 44) of the patients from Mainland China, the starting point of the collective response to the COVID-19 pandemic however remains unclear (*8*).

To analyze the collective response to COVID-19, digital information turns out to be critical, efficient, and effective (*6*). For example, mobile phone applications (Apps) can be easily used to conduct contact tracing and notification upon case confirmation (*6*). With data collected from the mobile phone Apps, statistical analysis has been carried out for COVID-19-related studies (*9–11*), e.g., Baidu Migration (*12*). Further, search engines, e.g., Baidu (*13*), has been used to understand social responses to the pandemic from the keywords of massive search queries while making users well-informed during the outbreak of COVID-19. In addition, mobility data from Baidu Maps also demonstrates the collective response from the perspective of mobility (*14,15*), where we can compare the massive mobility traces under the pandemic with the regular patterns in past years, so as to sketch how populations move as a response to COVID-19. In summary, please refer to studies (*16–27*) for analyzing the collective response to COVID-19 with digital information.

In this work, we focus on the analysis of the collective response in the early stage of the pandemic using both search data and mobility data. Specifically, we exploit the search index data from Baidu search index and mobility traces from Baidu Maps, where we intend to explore the potential abnormal changes of collective behaviors in the second half of 2019. To achieve the goal, with the search index data, we sort candidate keywords based on the correlation between the search volume of the keyword and the number of confirmed cases of COVID-19, using two methods, i.e., Dynamic Time Warping (DTW) (*28, 29*) and Detrended Cross-Correlation Analysis method (DCCA) (*30*). Then, we perform the correlation analysis from 2014 to 2020 between any two of the four regions, i.e., Wuhan, Mainland China, the United States (US), and the whole world, using DCCA. From the correlation, we try to identify abnormal changes and analyze the corresponding search volume and keywords, where we have not found any abnormal change before December, 2019 compared to previous years. With mobility data from Baidu Maps, we analyze the trend of population mobility from 2018 to 2020. We normalize the population inflow and outflow from June, 2018 to March, 2020, and we analyze the inflow and outflow separately on an annual time scale, where we try to identify the starting point of the collective response to the pandemic and observe how population flows change before and after the outbreak of COVID-19. Our analysis using search data and mobility data reveals that the collective response to the COVID-19 pandemic started from December, 2019 in Mainland China.

Compared with the existing work (*14, 31, 32*), we aim at tracing back the starting point of COVID-19 and observing the collective response in such a critical moment through time-series analysis on search data and mobility data analysis with massive mobility traces. In terms of determining the starting moment of COVID-19, (*9–11*) infer the possible origin dates of COVID-19 using dynamical systems and the numbers of cases confirmed over time. Compared to these studies, our work revisits the early stage of the pandemic from the perspective collective response, using search data and mobility data collected from massive mobile users in a participatory fashion. To uncover the latent factors of epidemiological dynamics, (*7, 16–26*) fit parameterized models with the numbers of cases confirmed over time, and interpret the models through the physical and epidemiological meaning of every parameter. Compared to these studies, our work does not make any assumptions on the epidemiological dynamics while providing a model-free analysis on the search and mobility data. Instead, we simply track the collective response through ranking the frequent search keywords and detecting the changes in population inflow/outflow between Chinese cities, in a data-driven fashion. Finally, this work incorporates direct evidences through multi-modal data fusion, while existing studies (*33*) rely on inferences and predictions based on indirect measurements. For example, (*33*) estimated and compared the volumes of visits to hospitals in Wuhan in 2018 and 2019 using remote sensing data from satellites, then claimed abnormalities detected. We directly sample the volumes of visits to hospitals in Wuhan from 2018 to 2020 through tracking the massive mobility traces collected from Baidu Maps, and confirm that there was no abnormality of hospital visit volumes before December, 2019 in Wuhan.

## Search Index Analysis

Using search index data, we analyze correlations between the frequent search keywords, related to respiratory diseases, per day and the trends of confirmed cases using DTW and DCCA. Specifically, we first analyze the long-term search data from March, 2014 to March, 2020, in four regions, i.e., Wuhan (Baidu Search Index), Mainland China (Baidu Search Index), the US (Google Trends) and the world (Google Trends), where we observe the spike of search trends with keywords related to respiratory diseases during the outbreak. Afterwards, we present the analysis on the second wave of the COVID-19 pandemic from March, 2020 to July, 2020, in Beijing., where we observe consistent patterns appeared in the collective response of the first wave.

### Correlation Analysis based on DTW and DCCA

We collected three datasets: the first dataset contains 47 COVID-19-related keywords; the second dataset includes the search index (2014 - 2020) from Baidu search index for Wuhan and Mainland China, and Google Trends for the US and the whole world; the third dataset consists of the number of confirmed cases per day from COVID-19 statistics (*34*).

As both search index data and the number of confirmed cases are time-series data, i.e., each data item associated with a time stamp, we use DTW and DCCA as analytical tools. DTW is introduced to adjust the time stamps and to match sequences that are similar but out of phase (*35*). DCCA is based on detrended covariance to investigate power-law cross-correlations between different simultaneously recorded time-series (*30*). Both DTW and DCCA can analyze the time-series data. For every keyword, we estimate the correlation using DTW and DCCA between the everyday search index of the keyword and the number of confirmed cases per day during the pandemic, where we believe the top keywords with highest correlations refer to the most relevant evidences and the correlations estimated by DTW and DCCA confirm each other. We rank the keywords in four regions, i.e., Wuhan, Mainland China, the US, and the world, separately, where the common top five keywords are:

- “Covid,” “epidemic situation,” “mask,” “pneumonia,” and “sterilization” (based on DTW);
- “Covid,” “epidemic situation,” “mask,” “sterilization,” and ‘nucleic acid” (based on DCCA).

In addition, we find that COVID-19 relative symptoms (*33*), such as “diarrhea” (16^*th*^) and “cough” (20^*th*^), are included within the top keywords

To analyze the early stage of the COVID-19 epidemic, we study the correlation of the search index of the high-ranked keywords between any two of the four regions (Wuhan, China, the US, and the world), using DCCA for each year between 2014 and 2020. We take “mask,” as an example, as “mask” is high-ranked in all the four regions.

From the analysis, we can see that there is no significant change in the correlation among the four regions before December, 2019. The correlation patterns between Wuhan and China, and between the US and the world, are consistently significant. By the end of 2019, the search index shows that the search volumes in the US or all over the world spiked faster than those in Wuhan or China. It is since the end of 2019, or more particularly, since 2020, that the COVID-19-related search volumes have increased significantly, reaching peaks in January or February in Wuhan and China. The search index of the keywords also reveals that the early stage of the collective response to the COVID-19 pandemic began from the end of 2019, i.e., December of 2019.

From Figure 1a, we can see that the correlations among the four regions were consistently positive and did not change much between 2014 and 2019, except in 2015, where positive correlations suggest the synchronizing trends of search keywords in global-wise. However, the correlations between Wuhan and the world, Wuhan and the US, the world and China, and the US and China have significantly changed and have dropped to negative in 2020, while the overall trends of search index dramatically increased in the same time, compared with other years. It demonstrates the happening of massive events (i.e., COVID-19) which changes the common trends of search at global scale. Interestingly, prior to COVID-19, we found a short term in 2015 with negative correlations between Wuhan and the world, Wuhan and the US, the world and China, and the US and China. In that year, an outbreak of Middle East Respiratory Syndrome (MERS) was found in Korea (*36*) and influenced several Asian countries. Similar patterns repeat in the status quo of 2020.

**Figure 1:**
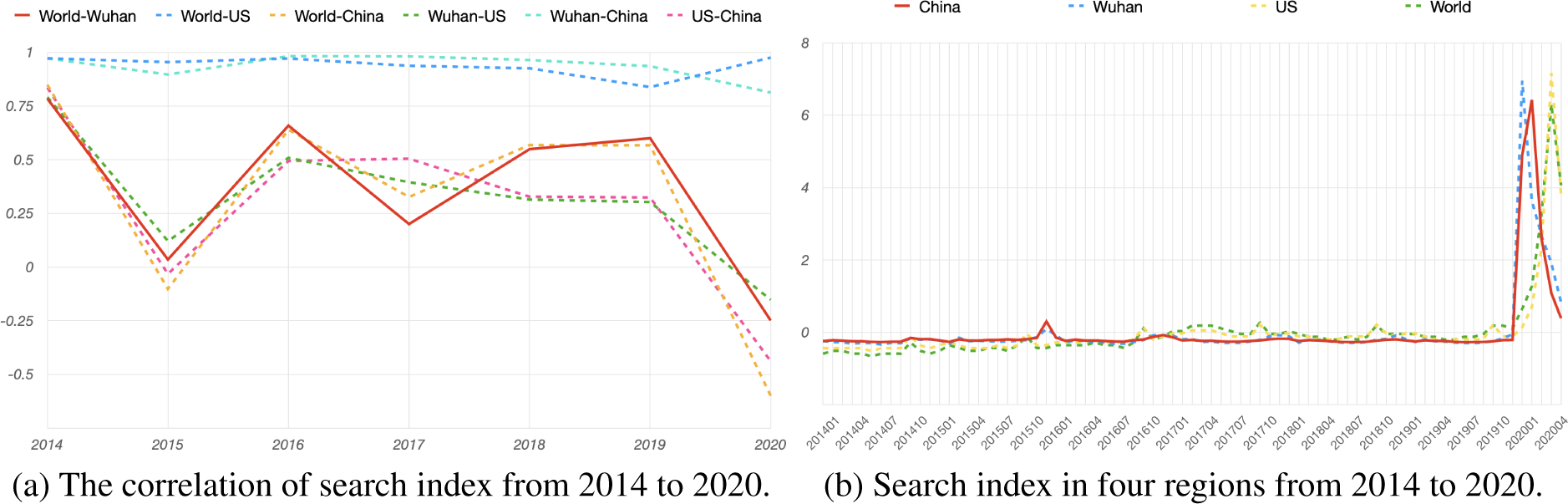
Analysis of search index from 2014 to 2020.

From search volumes shown in Figure 1b, we find that the search volumes in Wuhan or China before November, 2019 were still relatively lower than those in the US or the world. This result indicates that there was no special mask-related epidemic in Wuhan or China before November, 2019. In addition, we also find that the search volume of “mask” increased from December, 2019 and reached its peak in January, 2020 in Wuhan and China.

### Search Data Analysis of the Second Wave in Beijing

After great efforts were put into inhibiting the spread of COVID-19, the pandemic was mostly under control in China after May, 2020. However, on June 11th, the first case related to Xinfadi Market in Beijing was found, which led to a second wave of pandemic in Beijing. This second wave of epidemic persisted for more than a month, and was contained around the end of July. With previous experience in tackling COVID-19, this second wave was met with a swift response, not only in news reportage but also in measure implementation. We investigate how the public reacted to this resurgence of COVID-19 throughout this entire period by searching frequencies of keywords, and find that Beijing’s case is consistent with the previous national scenario.

From the analysis, we can see that the general public in Beijing exhibited a high level of attention to the overall event as the search volume of a majority of keywords, e.g., “nucleic acid,” “epidemic,” “mask,” “sore throat,” and “fever,” surges significantly following the outbreak on June 11th, where the search index of “nucleic acid” shown in Figure 2 is an example. However, a few words, e.g., “stuffy nose,” “respiratory failure,” and “respiratory distress,” which are mostly related to critically ill patients, did not show a noticeable change in search, which is in accordance with the fact that although the outbreak aroused concerns at first, quick containment actions prevented the outbreak from resulting in a huge number of significantly ill patients. Until July, the search trends fell back into the previous levels as normalized in earlier months, since the epidemic was fully under control after mid-July.

**Figure 2:**
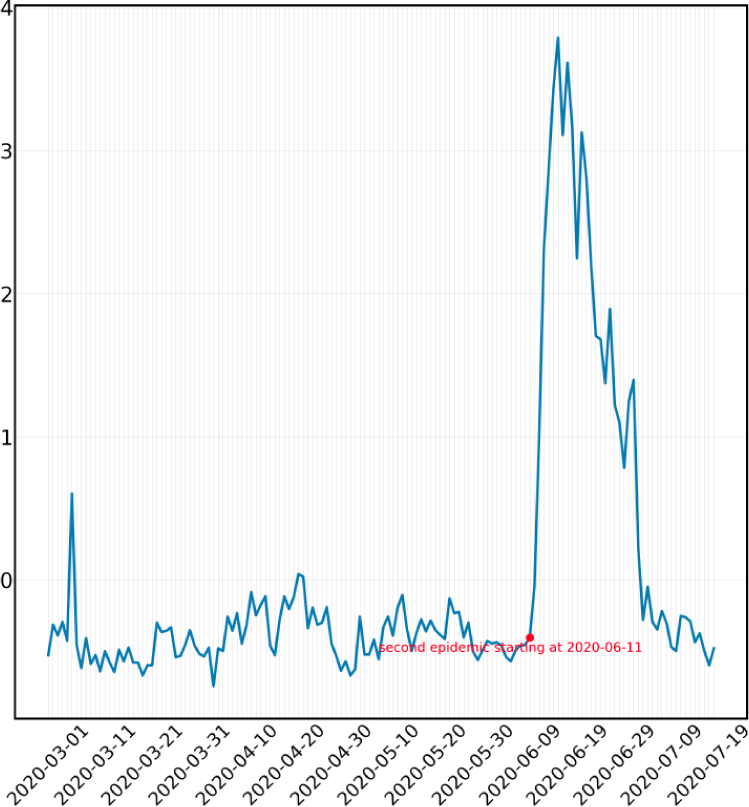
Search index of “nucleic acid” in Beijing from March 1st, 2020 to July 20th, 2020

## Mobility Traces Analysis

In this section, we try to find abnormal population mobility trends from June, 2018 to March, 2020, exploring the starting point of the collective response to COVID-19. We exploit the mobility data from Baidu Maps, which consists of the inflow and outflow to/from Wuhan, and the population densities of multiple hospitals in Wuhan, from June, 2018 to March, 2020.

### Inflow and Outflow Analysis

Figures 3 and 4 show the population inflow and outflow in Wuhan, spanning from June, 2018 to March, 2020, extracted from the mobility traces from Baidu Maps. Each point of data representing inflow refers to the number of people who entered into the city on that specific date, and correspondingly, each outflow data point refers to the number of people who left the city on that date. We separate inflow and outflow into distinct diagrams in order to demonstrate a sharper focus on time scale comparison. In addition, all absolute values of the mobility data were standardized to shed light on a more structured inference on an equal baseline across different times. For a few dates with missing data, we utilize the data from a previous date as an estimation.

**Figure 3:**
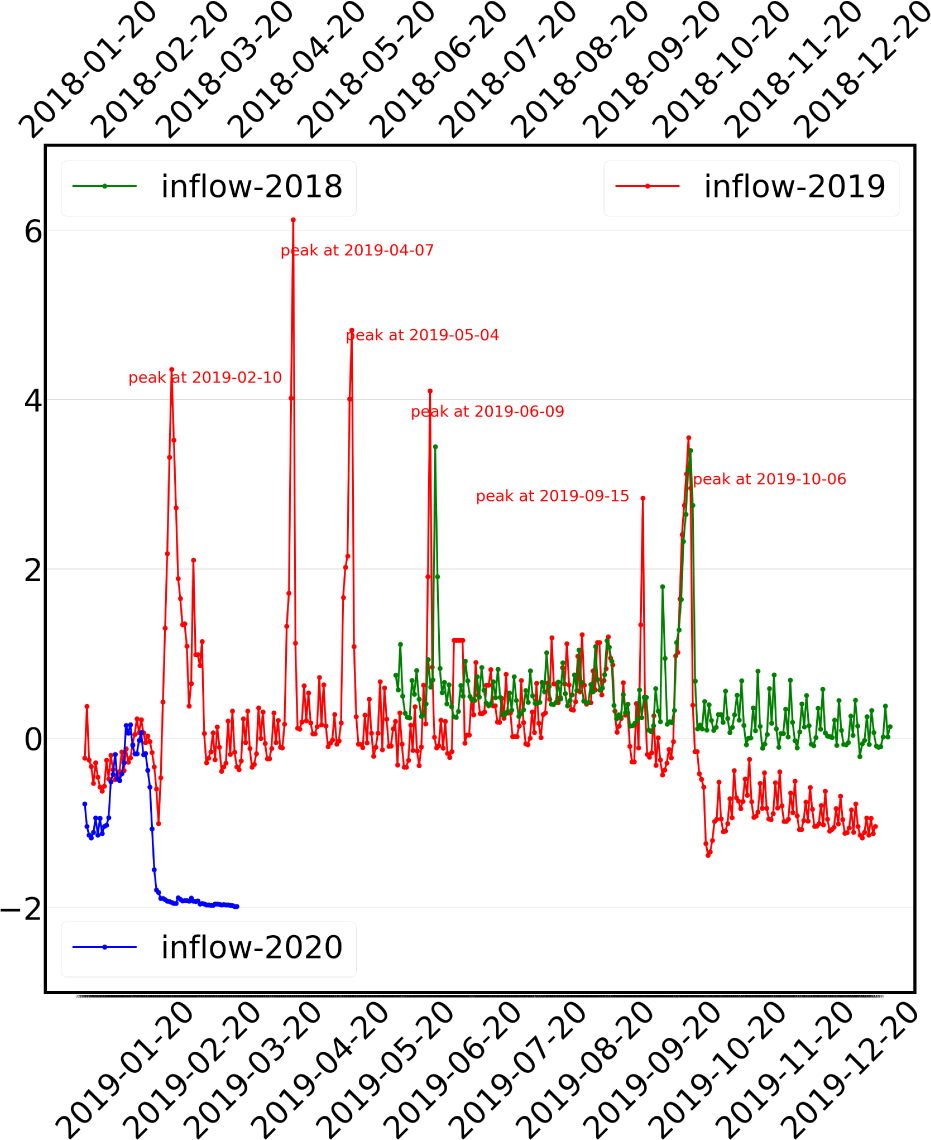
The population inflow into Wuhan (2018 - 2020). The inflow of 2019 is between January 1st and December 31st. The inflow of 2018 is between June 1st and December 31st, which is aligned with that of 2019 using the Solar Calendar. The inflow of 2020 is between December 22nd, 2019 to March 31st, 2020, which is aligned with that of 2019 using the Lunar Calendar.

**Figure 4:**
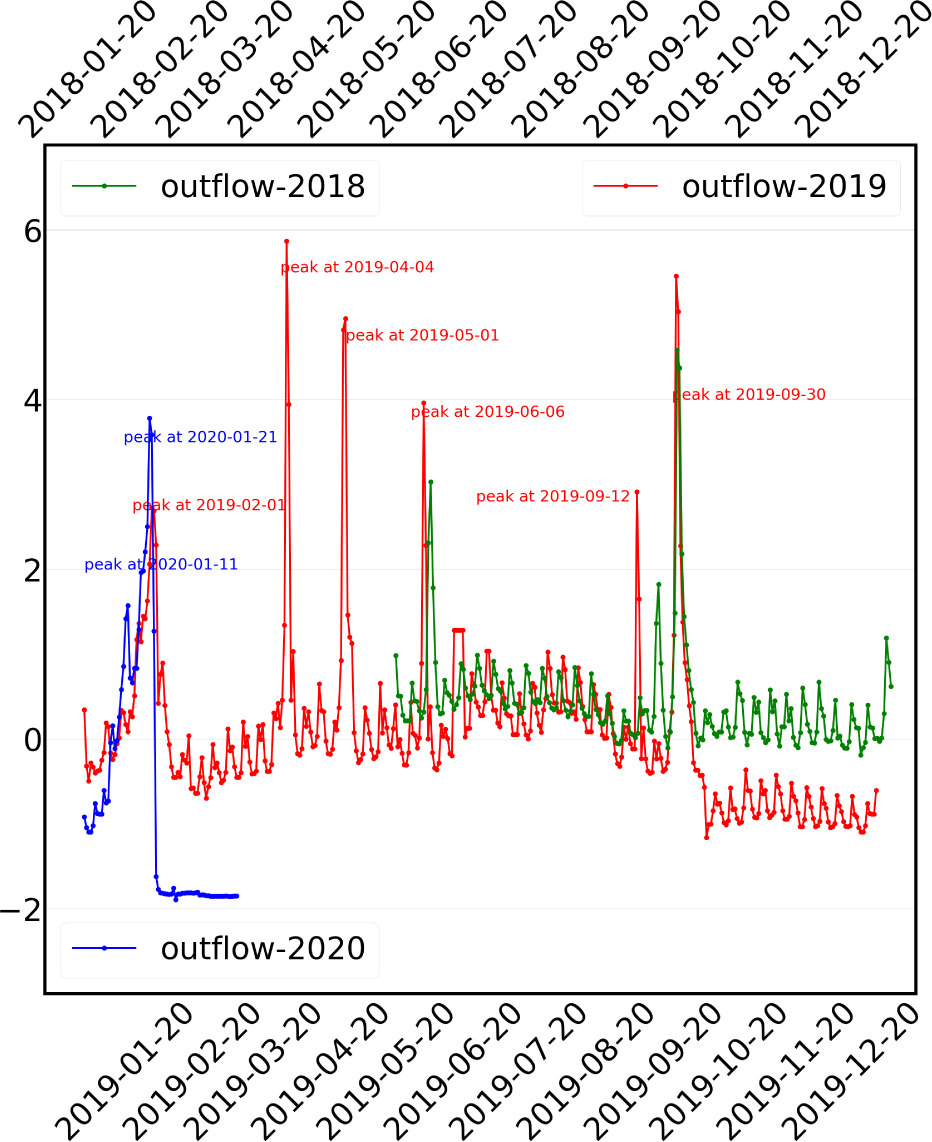
The population outflow from Wuhan (2018 - 2020). The inflow of 2019 is between January 1st and December 31st. The inflow of 2018 is between June 1st and December 31st, which is aligned with that of 2019 using the Solar Calendar. The inflow of 2020 is between December 22nd, 2019 to March 31st, 2020, which is aligned with that of 2019 using the Lunar Calendar.

#### Analysis of Inflow

As shown in Figure 3, we find that the collective mobility related response to the pandemic has happened around January 24th (Spring Festival), 2020. For inflow data shown in Figure 3, we align inflow traces from January 1st, 2018 to December 31st, 2018 along with January 1st, 2019. We obtain data starting from June 1st, 2018, and thus the green line representing the inflow of 2018 started from the second half of the figure. As we can see from the figure, these two lines of 2018 and 2019 exhibit a high level of superposition, especially similar pattern trends in corresponding time spots for peaks and troughs. The peaks that happened around June 9th, September 15th, and October 6th were mainly due to Chinese national holidays, including the Dragon Boat Festival, the Mid-Autumn Festival, and National Day, where people flooded in for family gatherings, travel, or other leisure purposes. The 2018 line displays a stable trend bounded around 0 and 1 throughout the second half of the year, while the line of 2019 shows a slight decline in inflow after the National Day holiday, compared to previous parts of the year. Yet, the overall trend is steady, without significant stumbling. The line representing inflow of 2020 contains the inflows from December 22nd, 2019 to March, 2020. Note that in Figure 3, we align the yearly traces according to the Lunar Calendar for better comparison. We can see both peaks happened around Spring Festival due to the national holidays. It is after Spring Festival that we witness a significant decline in inflow approaching a standardized score of −2, as the pandemic began and China announced travel restrictions. From the population inflow traces, we conclude that there is no obvious abnormal trend before Spring Festival in Wuhan compared to previous year’s data, which implies our finding.

#### Analysis of Outflow

As shown in Figure 4, with the analysis of outflow, we also find that the collective mobility related response to the pandemic has happened around Spring Festival. In Figure 4, two lines of 2018 and 2019 exhibit a high level of superposition, with peaks around national holidays similar to scenarios in inflow diagrams. Zooming in for the latter part of 2019, the outflows from October to December maintain a paralleling trend, indicating no aberrant leaving or public panic. The increasing slopes of the outflow are similar in 2019 and 2020, as Wuhan is a city with abundant migrant workers. Yet, we can see some abnormal humps in the outflow at January 11th, 2020, where outflow is obviously above the previous year’s level, probably suggesting an onset of COVID-19 and more people leaving the city to avoid it. It is after Spring Festival that we witness a significant decline in outflow, approaching a standardized score approximating −2, implying that the start of the collective response to the pandemic is likely to have happened around Spring Festival.

Finally, the combination of the analysis of inflow and outflow further confirms that the collective mobility related response to the pandemic have happened three days before Spring Festival, i.e., January 21st, 2020. For speculating a point as a start of the collective response to the pandemic, we need to find abnormal trends with high outflow and low inflow. If people were aware of abnormal events, people within the city would want to leave and people outside of city would not want to enter. Yet, we do not note any significant declining trend in inflow nor rising trend in outflow during the second half of the year in 2019. We find an abnormal peak of outflow around January 11th, 2020, and a higher increase in outflow than the previous year, peaking around January 21st, 2020. Reflected within the data, we also see a highly efficient government measure implementation, as both inflow and outflow tumbled after travel restrictions (from/to Wuhan) announced on January 23rd, 2020, hitting an unprecedented level of almost −2, and this low level persisted afterwards.

#### Analysis of Population in the Hospitals of Wuhan

Figure 5 shows the normalized total population in 26 hospitals of Wuhan, including Tianyou Hospital affiliated to the Wuhan University of Science and Technology mentioned in (*33*), which have accepted COVID-19 patients. Each point of data represents the population index at that specific category of locations on each date spanning from June, 2018 to March, 2020, based on data from Baidu Maps. All absolute values of the mobility data were normalized to shed light on a more structured inference on time scale comparison. For days that we miss, we use data from a previous date as an estimation. Especially for 2018, where we may observe from the exceptionally parallel and ladder-like curve, we are missing some parts of data.

**Figure 5:**
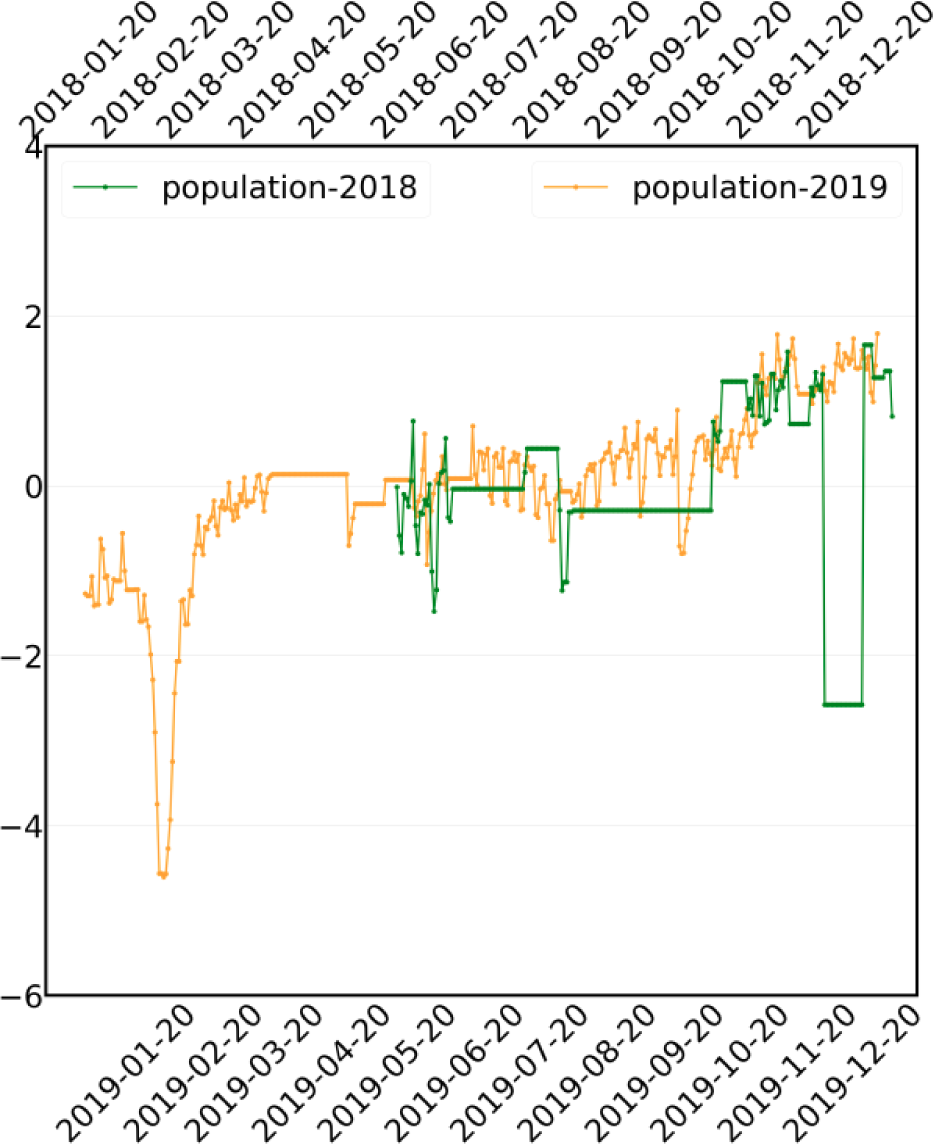
Population data of hospitals in Wuhan 2018-2019.

As well, the analysis of the population in the hospitals of Wuhan suggests that the collective response did not start before the end of 2019. By concentrating on the figures of hospitals, we align the data based on years. It was suspected that COVID-19 infections had begun at the second half of the year (*33*), so we mainly focus on the latter part of this timeline. The two lines of 2018 and 2019 were matched in similar trends. Generally, we see a nearly parallel shape from June to mid-September. It is after mid-September that we see a discernable climbing trend of population visiting hospitals, and the higher level persists as a plateau shape until the end of the year. Despite considerable missing parts of 2018 data, we can still see a jump between mid-year average level and end-of-year average population level as population visiting hospitals may rise due to seasonal reasons, e.g., people are more contagious and vulnerable to diseases when temperatures fall. Yet, we can see that the gap between the peak levels happening in the end of 2019 and the peak levels happening in the end of 2018 remains almost unchanged, as in early periods in the year. Thus, the trend did not show significantly abnormal change in late 2019. This phenomenon greatly reduces the strength of any suspicion that the collective response to the pandemic might have begun in fall of 2019 (*33*), as the number of people visiting hospitals did not witness any aberrant rise in trends compared to the previous year.

## Conclusion

Search index is an important indicator for monitoring the outbreak of a pandemic. In this work, we analyzed the correlation of search index corresponding to COVID-19-related keywords. We first ranked the keywords based on the correlation of the search index and the number of confirmed cases of COVID-19. We found that “mask,” “cough,” and “diarrhea” are within the 28 highly ranked keywords but not within the top rankings among all. Then, we analyzed the correlation between any two regions among the four regions (Wuhan, Mainland China, the US, and the world). We found that the correlations and the search index did not change much (because of COVID-19) before December, 2019, in Wuhan or in Mainland China. We thus argue that the collective activities start from December, 2019 in Mainland China.

Furthermore, we analyzed the mobility data of Wuhan and the population in 26 hospitals of Wuhan to confirm our claim. By comparing the trends in the second half of 2019 with that of 2018, we did not find abnormal deviations from the previous year, indicating that the start of the collective response to the pandemic occurred after December, 2019. Additionally, the increasing outflow and decreasing inflow, which signaled the pandemic, was not observed in the data of Baidu migration in Wuhan in the second half of 2019, further supporting the claim.

## Data Availability

While the COVID-19 outbreak is making an impact at a global scale, the collective response to the pandemic becomes the key to analyzing past situations, evaluating current measures, and formulating future predictions. In this paper, we analyze the public reactions to the pandemic using search engine data and mobility data from Baidu Search and Baidu Maps respectively, where we particularly pay attentions to the early stage of pandemics and find early signals from the collective response to COVID-19. First, we correlate the number of confirmed cases per day to daily search queries of a large number of keywords through Dynamic Time Warping (DTW) and Detrended Cross-Correlation Analysis (DCCA), where the keywords top in the most critical days are believed the most relevant to the pandemic. We then categorize the ranking lists of keywords according to the specific regions of the search, such as Wuhan, Mainland China, the USA, and the whole world. Through the analysis on search, we succeed in identifying COVID-19 related collective response would not be earlier than the end of 2019 in Mainland China. Finally, we confirm this observation again using human mobility data, where we specifically compare the massive mobility traces, including the real-time population densities inside key hospitals and inter-city travels departing from/arriving in Wuhan, from 2018 to 2020. No significant changes have been witnessed before December, 2019. Our paper is based on the Baidu index, google trends and the mobility data from Baidu Huiyan.

http://index.baidu.com/v2/index.html#/

https://trends.google.com/trends/?geo=US

https://huiyan.baidu.com/

## Acknowledgements

All experiments in this paper were carried out using anonymous data and secure data analytics provided by Baidu Data Federation Platform (Baidu FedCube). For data accesses and usages, check http://fedcube.baidu.com/page_en.html.

## Supplementary Materials

In this section, we present the details on data normalization, correlation analysis of other keywords, and some other details.

### Data Normalization

To visualize the search index data of keywords in different regions in a single figure, we plotted figures using normalized data. In Figures 1, S1, S2, S3, S4, S5, S6,S7, S8, S9, S10, S11, S14 and S15, we visualized the search index data of different keywords in four regions, i.e., Wuhan, China, the US, and the world. Given the search index of a keyword in a region from January, 2014 to June, 2020, we normalized the data using *Z-score standardization*. Given the overall volumes of the search volume in a region from January, 2014 to June, 2020, we first calculated the mean value and the standard derivation value of the search index as *µ* and *σ* respectively. Then we normalized the search index in each region as

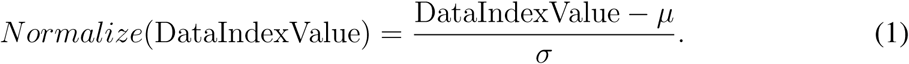

In this way, the search index of each keyword in the study is normalized to almost the same scale, which fits into the same figure. Similarly, data normalization was also applied when we calculated the inflow, outflow and the population in the hospitals of Wuhan from June, 2018 to March, 2020.

### Ranking Results

In this section, we present the detailed ranking results using DTW and DCCA. Table S1 represents the ranking of the top 28 keywords in Wuhan using DTW and DCCA. From the table, we can see that the common highly (top 10) ranked keywords are “COVID,” “Epidemic situation,” “Nucleic acid,” “Pneumonia,” “Sterilization,” and “Infection” based on DTW and DCCA. In addition, “Mask” is highly ranked (4^*th*^) using DTW. Table S2 represents the ranking of the top 28 keywords in China using DTW and DCCA. The common highly ranked keywords are “COVID,” “Nucleic acid,” “Runny nose,” “Sterilization,” and “Hypoxemia” based on DTW and DCCA. In addition, “Mask” is also highly ranked using DTW (8^*th*^) and DCCA (12^*th*^). Table S3 demonstrates the ranking of the top 28 keywords in the US using DTW and DCCA. The common highly ranked keywords are “COVID,” “Mask,” “Sterilization,” “Body temperature,” and “Epidemic” based on DTW and DCCA. “Mask” is also highly ranked using both DTW (2^*nd*^) and DCCA (1^*st*^). Table S4 demonstrates the ranking of the top 28 keywords all over the world, using DTW and DCCA. The common highly ranked keywords are “COVID,” “Mask,” “Sterilization,” “Body temperature,” “Epidemic situation,” “Respiratory distress,” and “Respiratory failure” based on DTW and DCCA. “Mask” is highly ranked using both DTW (2^*nd*^) and DCCA (2^*nd*^).

### Correlation Analysis of “Cough” and “Diarrhea”

In this section, we present the correlation analysis on “Mask,” “Cough,” and “Diarrhea”. “Cough” and “Diarrhea” are considered as key symptoms for COVID-19 in (*33*).

#### Correlation Analysis of “Mask”

The search volumes of “mask” in four regions are shown in Figures S1 and S2.

#### Correlation Analysis of “Cough”

Similar to “mask,” as shown in Figure S3, we can see that the correlations were positive and did not change much between 2014 and 2019. In 2015, the correlations became less significant, which may also have been caused by the outbreak of Middle East Respiratory Syndrome (MERS), as “cough” is one of the most important symptoms (*36*). However, the correlations have changed to be negative in 2020. In addition, the search index also significantly increased in 2020 in Wuhan and China, which indicates the collective response to the COVID-19 pandemic in 2020.

**Figure S1:**
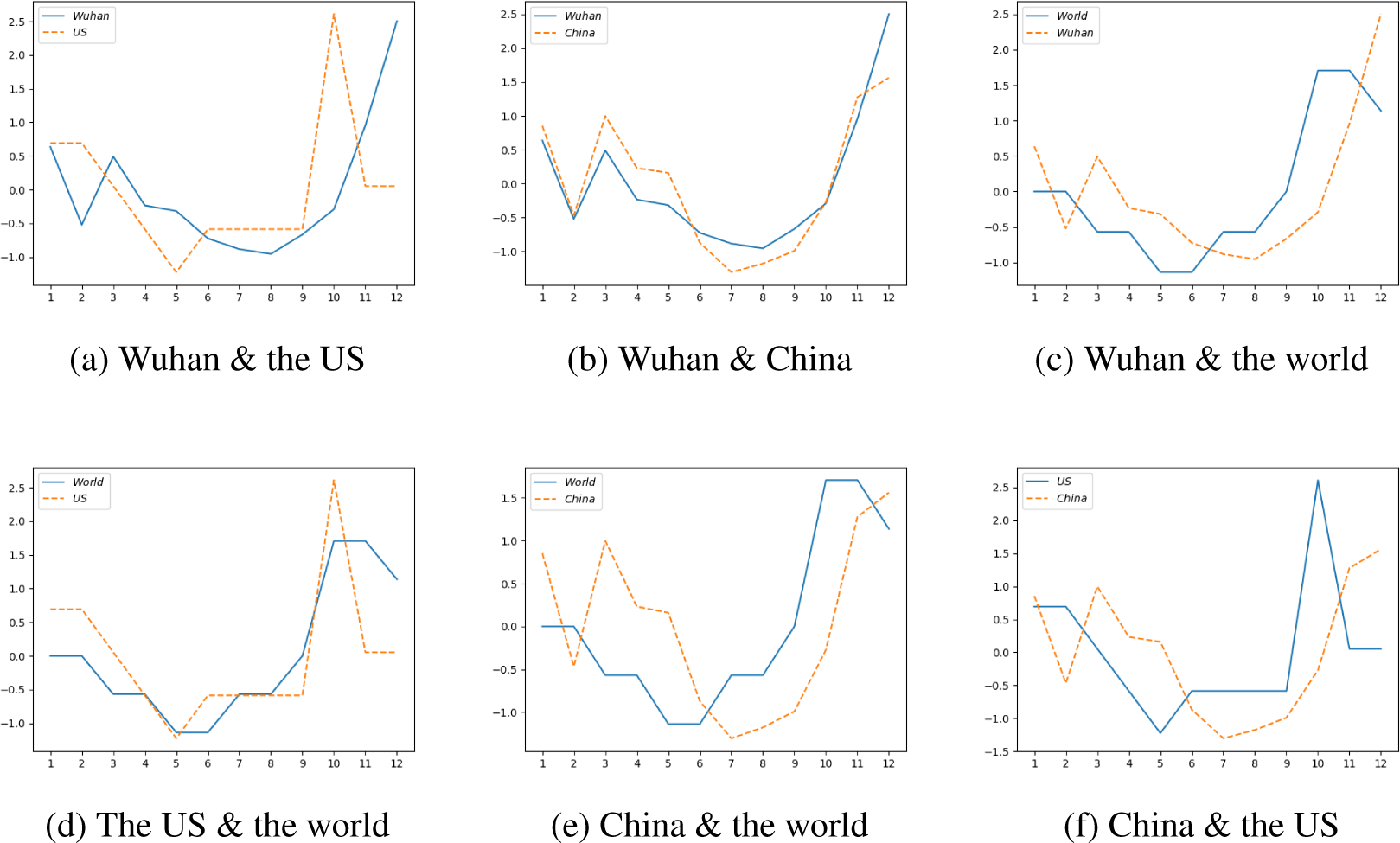
Comparison of the search volume of “mask” between two regions in 2019 with normalized data.

To analyze the early stage of the outbreak of COVID-19, we zoom in on 2019 and 2020. Figure S4 shows that the search volumes increased much faster in US or World than in Wuhan or China from August, 2019 to December, 2019. This result indicates that there was no evidence to support the outbreak of cough-related epidemic in Wuhan or China before December, 2019, compared with the US and the whole world. In addition, Figure S5 shows that the search volumes of “cough” increased from November, 2019 and reached the peak in January, 2020 in Wuhan and China. The volumes spiked significantly in Wuhan and China in December, 2019 compared with the US and the world. This is due to the fact that early confirmed cases were found in December, 2019 (*3, 37*).

**Figure S2:**
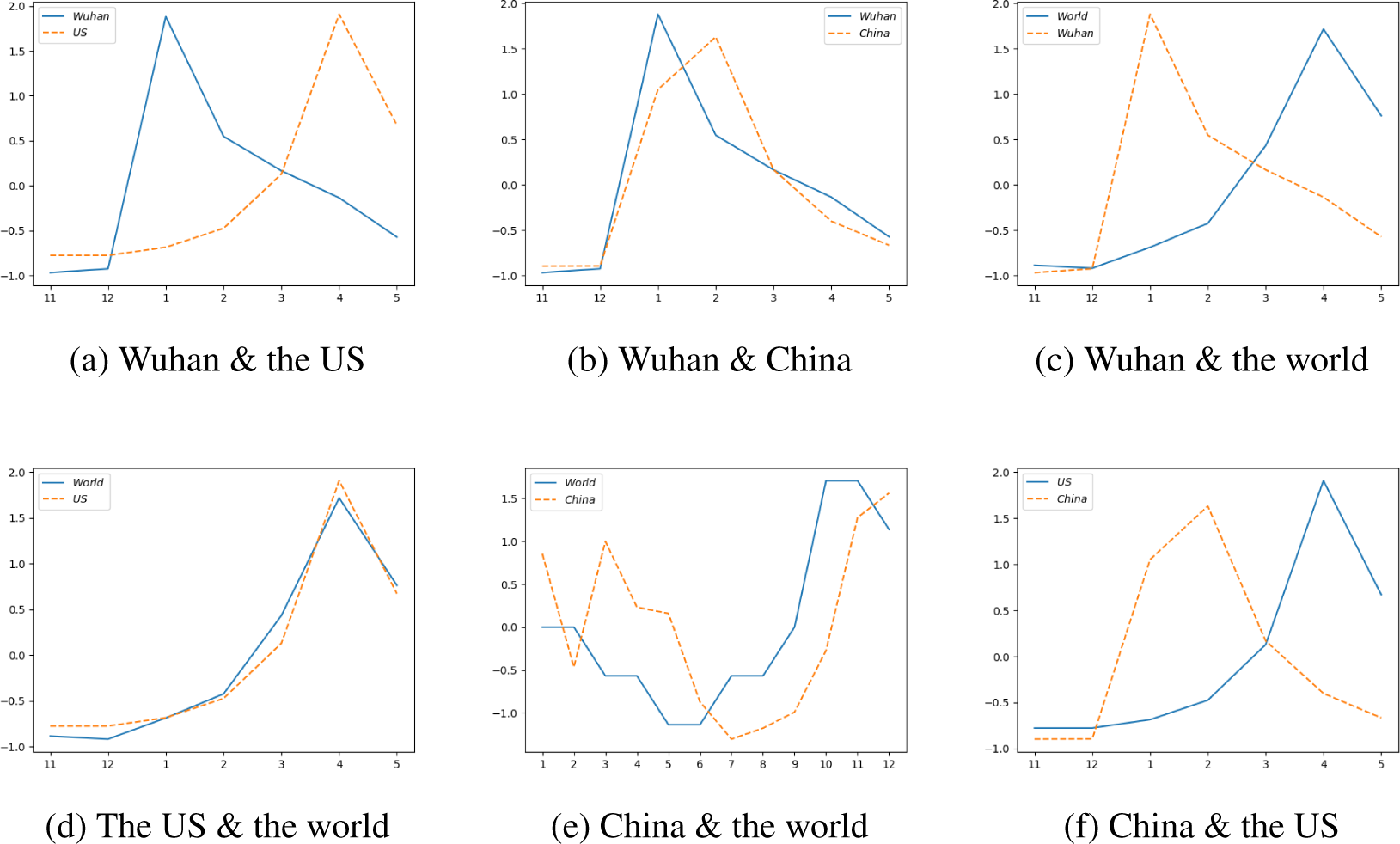
Comparison of the search volume of “mask” between two regions in 2020 with normalized data.

**Figure S3:**
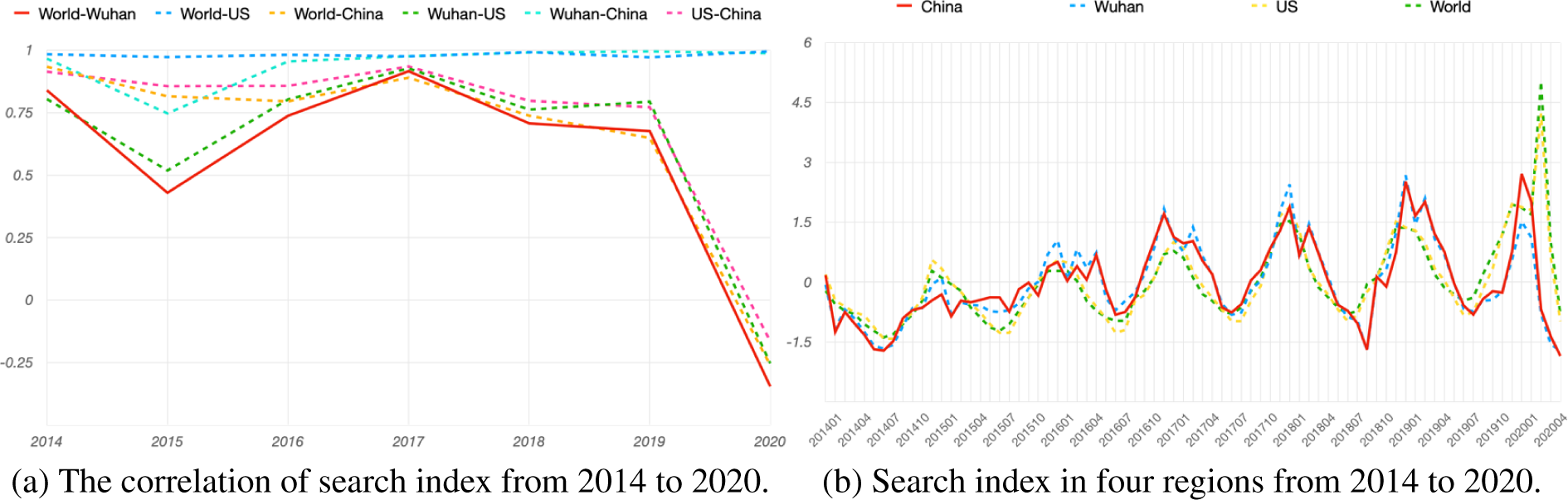
Analysis of search index of “Cough” from 2014 to 2020.

**Figure S4:**
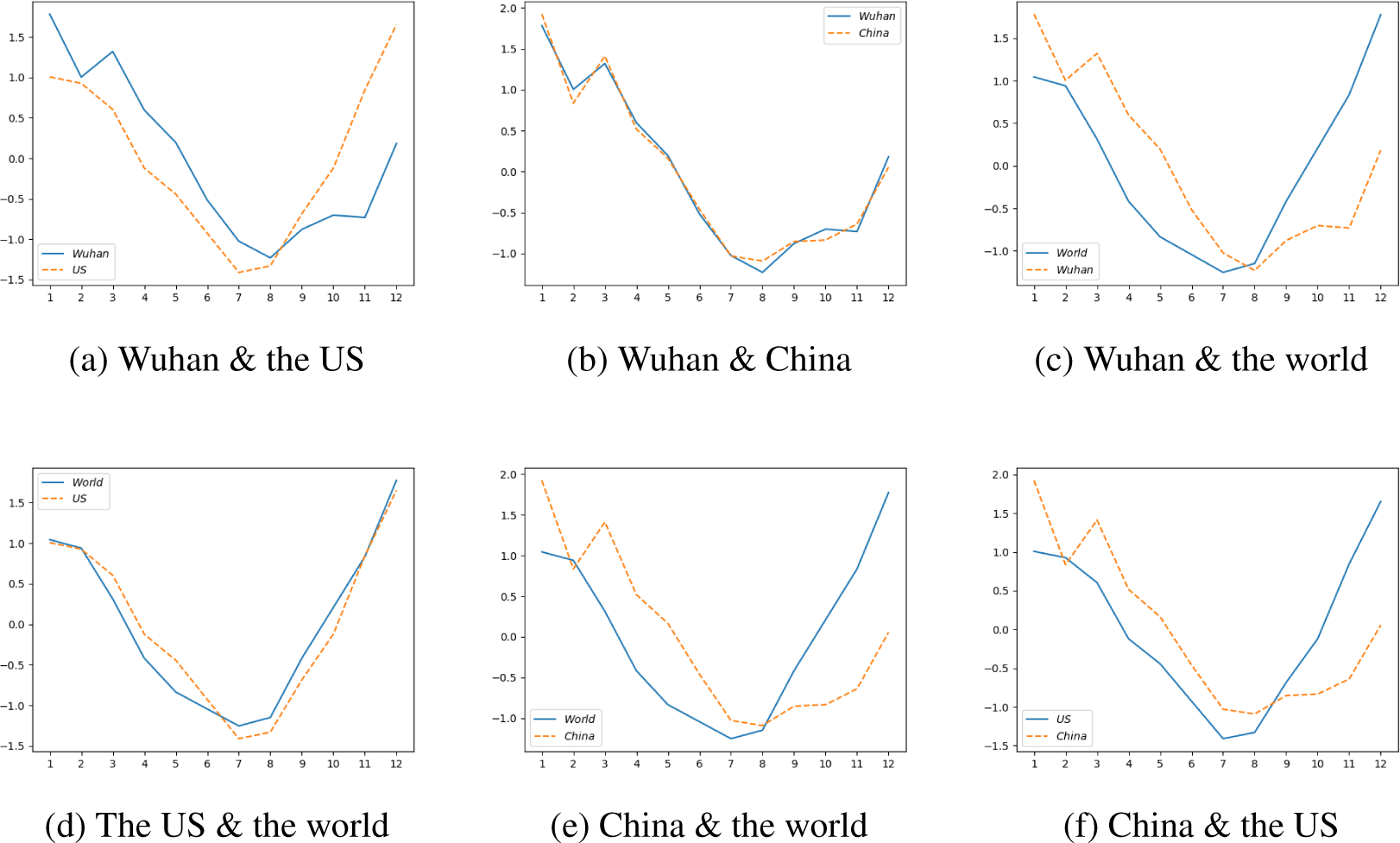
Comparison of the search volume of “cough” between two regions in 2019 with normalized data.

#### Correlation Analysis of “Diarrhea”

The correlation analysis of “Diarrhea” is shown in Figure S6, we can see that the correlation stayed positive and the search index increased from 2014 to 2018. In 2019, the correlations became a little bit less significant. In addition, the correlations and the search index (in Wuhan and China) have dropped to negative in 2020, which indicates the collective response to the COVID-19 pandemic in 2020.

To analyze the decrease of the correlation from 2019, we zoom in on 2019 and 2020. Figure S7 shows that the search volumes actually decreased in Wuhan or China from June, 2019 to November, 2019. This result indicates that there was no evidence to support the outbreak of diarrhea-related epidemic in Wuhan or China before December, 2019. In addition, Figure S8 shows that the search volume of “diarrhea” increased from December, 2019 and reached its peak in January, 2020 in Wuhan and in February, 2020 in China. Similar to the “cough,” the search volumes of “Diarrhea” spiked in Wuhan and China in December, 2019 compared to the US and the world. This is also due to the fact that early confirmed cases were found in December, 2019 (*3, 37*).

**Figure S5:**
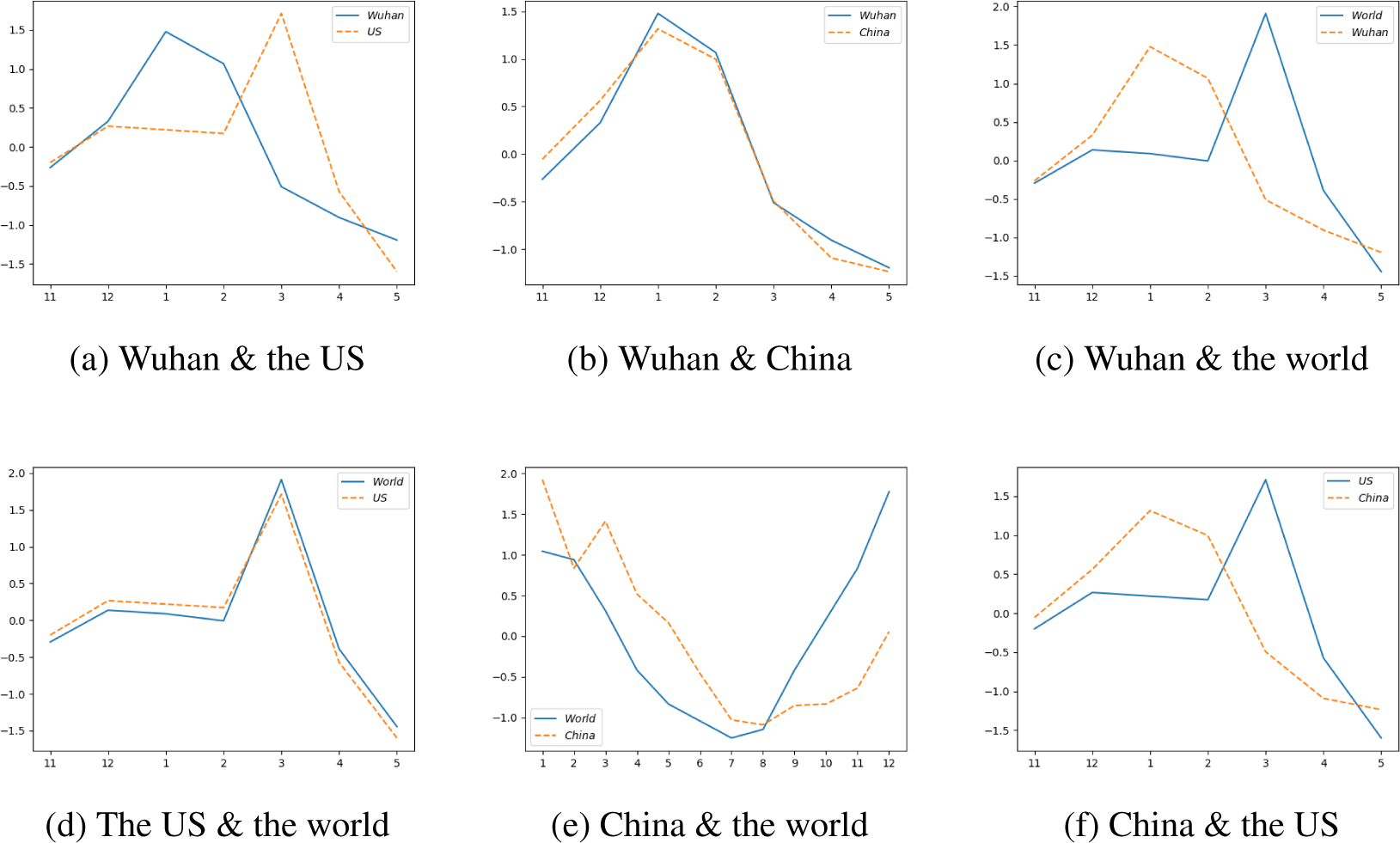
Comparison of the search volume of “cough” between two regions in 2020 with normalized data.

**Figure S6:**
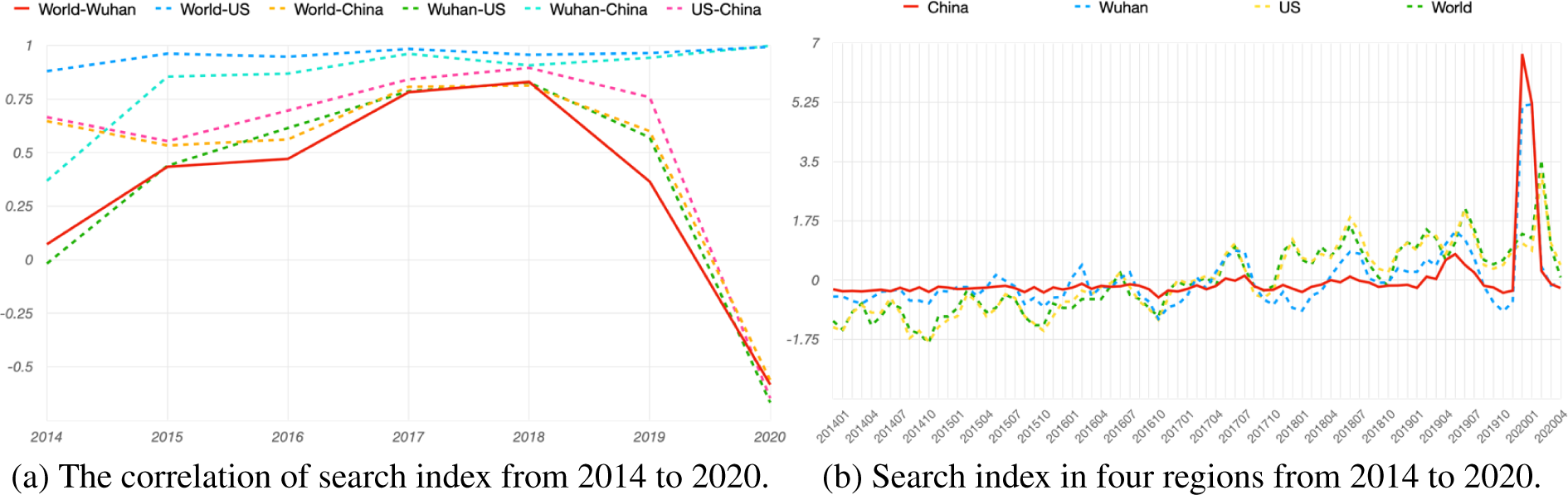
Analysis of search index of “Diarrhea” from 2014 to 2020.

**Figure S7:**
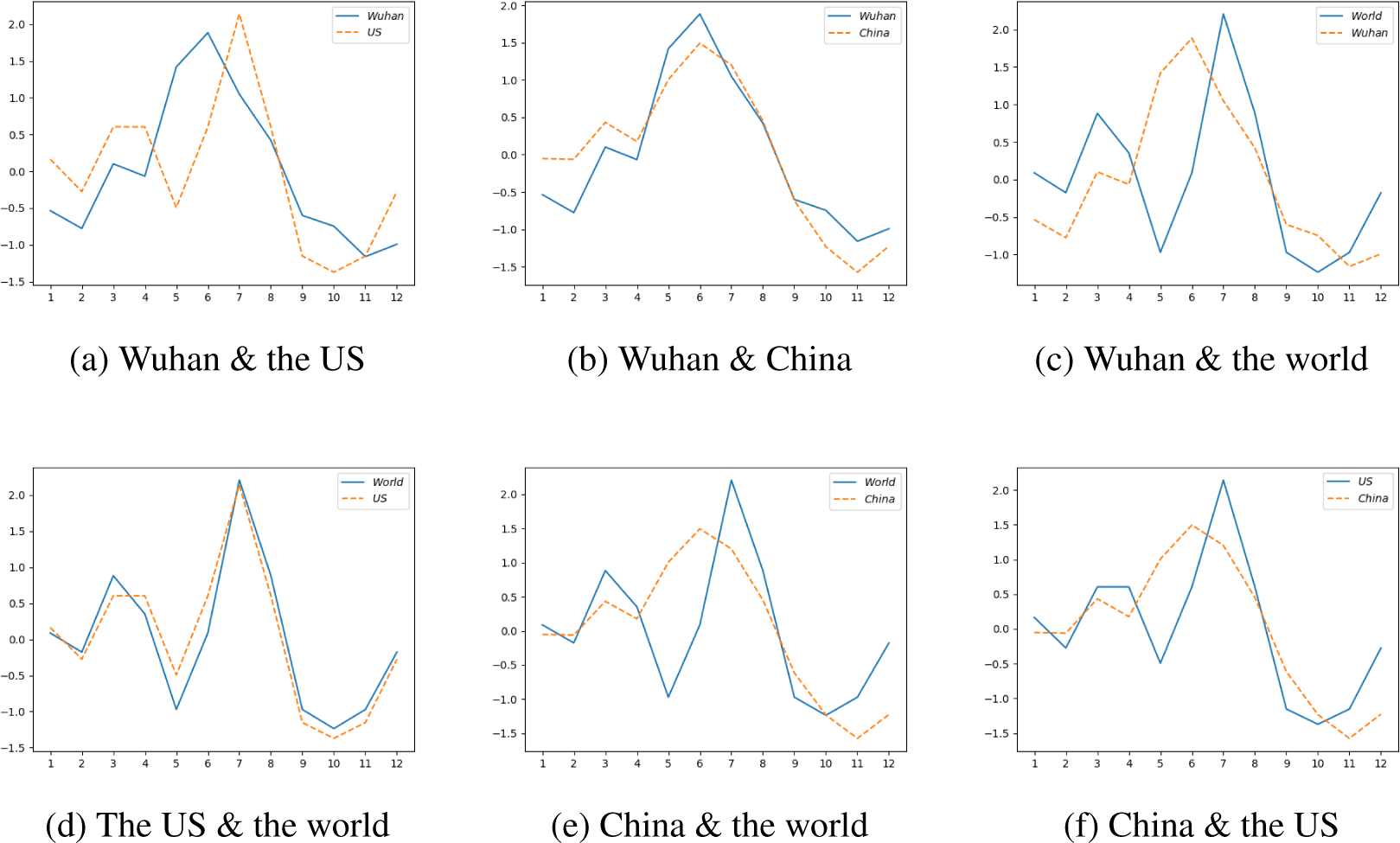
Comparison of the search volume of “diarrhea” between two regions in 2019 with normalized data.

#### Changes of Correlation Coefficients Over Time

In Figures S9, S10, and S11, we visualize correlations using DCCA and the search index in four regions for 3 other keywords. The correlations were positive and did not change much before 2020 except 2015 because of MERS. The correlations have significantly decreased in 2020, which coincided with the outbreak of COVID-19 in 2020. In addition, the search index of the 3 keywords reveals that the early stage of the collective response to the COVID-19 pandemic starts from the end of 2019, i.e., December of 2019.

**Figure S8:**
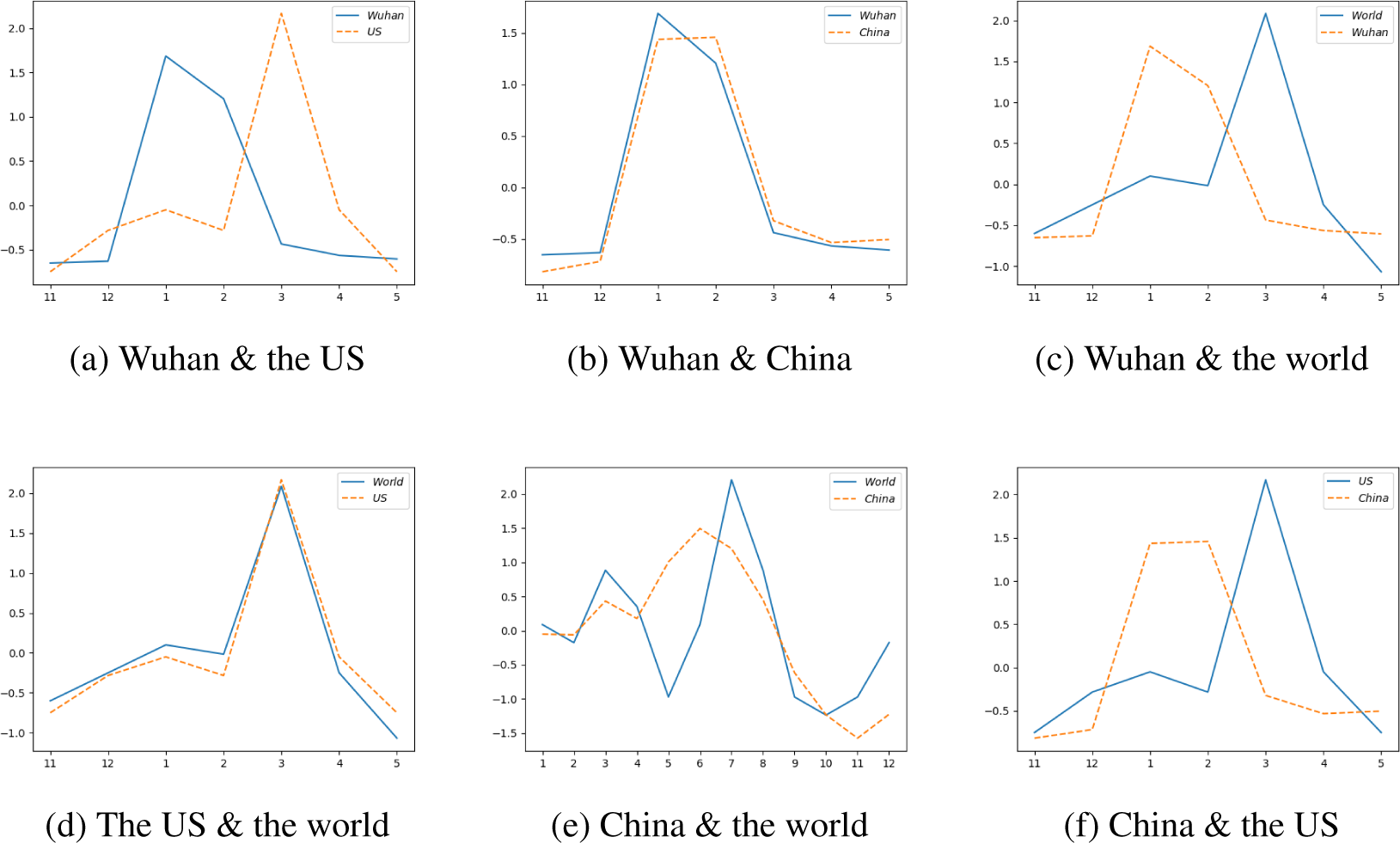
Comparison of the search volume of “diarrhea” between two regions in 2020 with normalized data.

**Figure S9:**
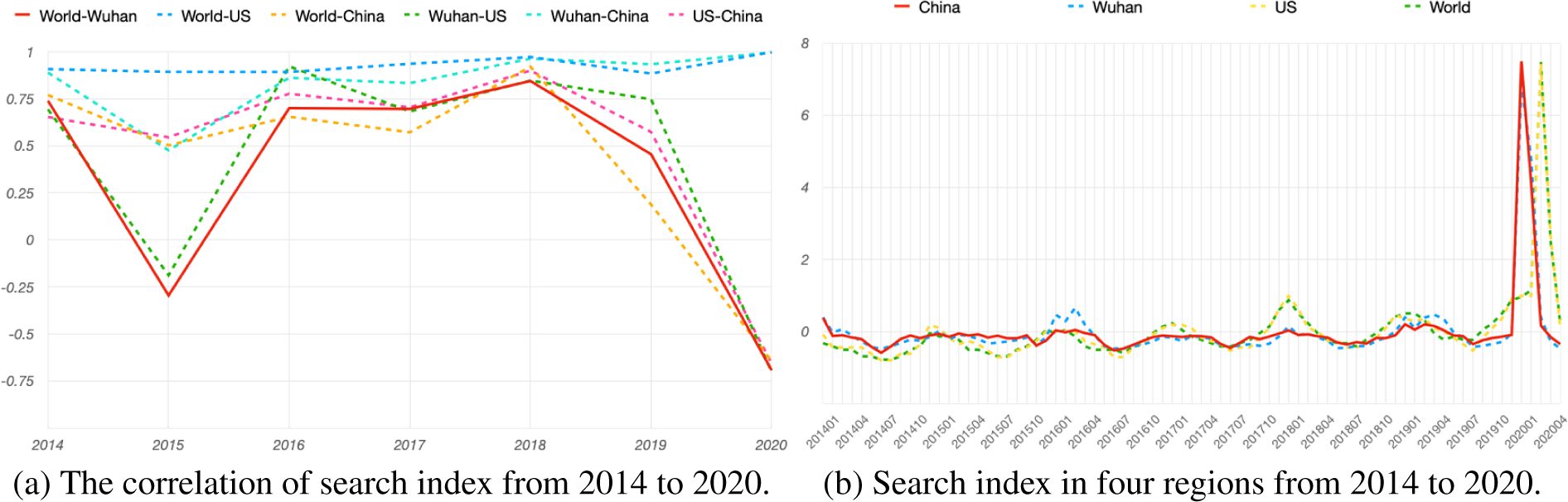
Analysis of search index of “dry cough” from 2014 to 2020. The decrease in 2015 is likely to have been caused by the outbreak of MERS in Korea.

**Figure S10:**
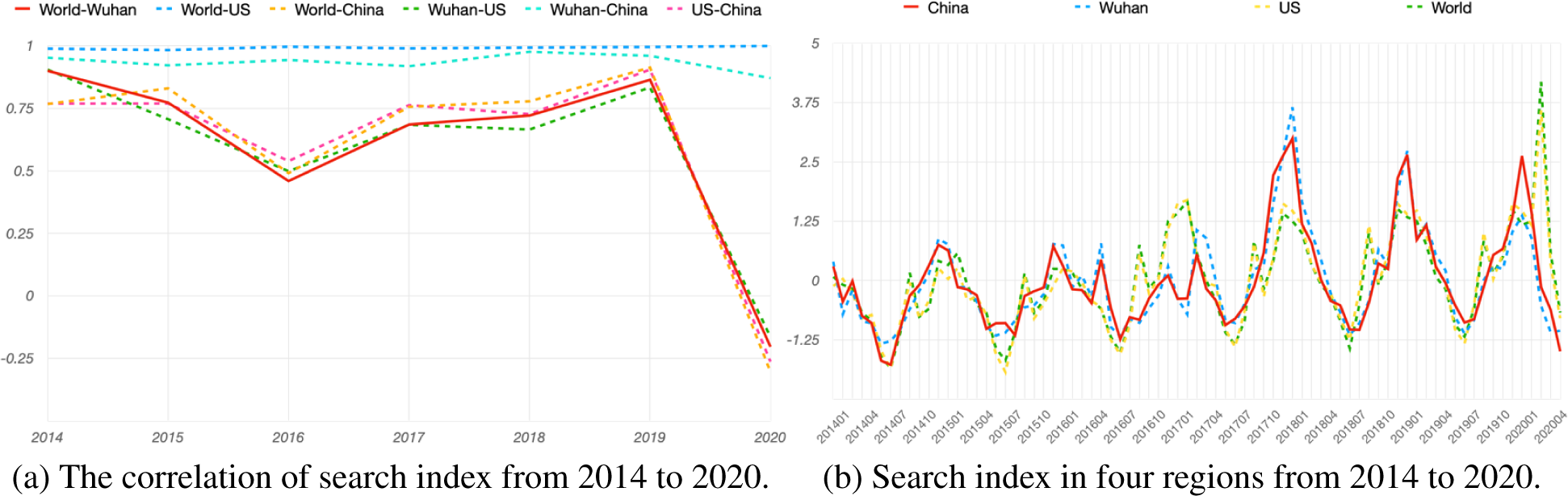
Analysis of search index of “stuffy nose” from 2014 to 2020.

**Figure S11:**
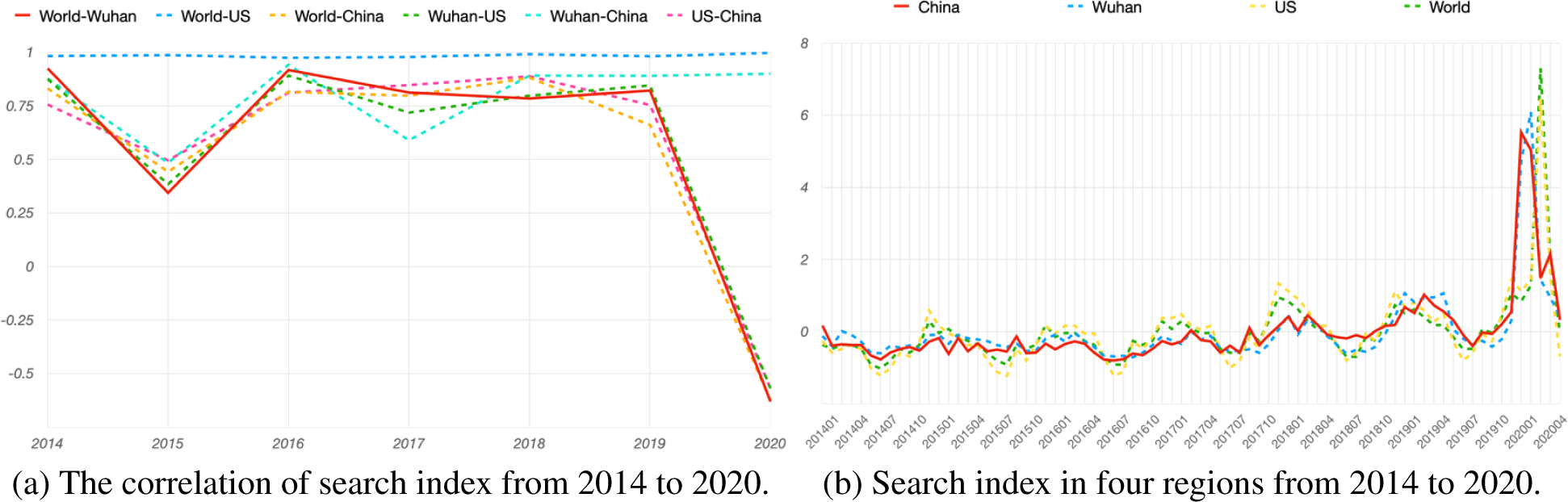
Analysis of search index of “sore throat” from 2014 to 2020.

#### Spearman’s Rank Correlation Analysis

We examined the correlations of search trends between every pairs of the four geographical regions, i.e., China, Wuhan, the US, and the world, using Spearman’s rank correlation (*38*).

With the correlation coefficients, we apply the hypothesis test of the “significance of the correlation coefficient” to decide whether the correlation between two search trends is significant or not. We use the significant level of 0.05, which is *α* = 0.05. Thus, we consider the correlation with p-value less than 0.05 as “significantly correlated” ; otherwise, the correlation is considered “insignificant.” By summing up the number of queries with significant correlated trends, an indicator for overall correlation of epidemic-relation search queries between two regions can be drawn.

To clearly identify the changing trends from 2014 to 2020, we separate the data into 7 subsets according to the corresponding year. Figure S12 shows the yearly trends for the number of significant queries between two of the four examined regions. Except for 2020, during which the number of queries with significant correlated trends between World and USA decreases to 18, the overall trend with high correlation can be observed. The decreasing number of queries with significant correlated trends can be summarized as that the status of the USA is different from the global trends in terms of the epidemic. The number of queries with significant correlated trends reveals an overall decreasing pattern from 2014 to 2020. Especially in 2020, all the queries we examined are insignificant, when comparing their global trends and the trends in China. Similar to the above comparison, the number of queries with significant correlated trends between World and Wuhan reveals an overall decreasing trend from 2014 to 2020. In 2020, the number of queries with significant correlated trends decreases to 2. This matches the outbreak of COVID-19 in Wuhan. In 2020, only the search trends of 2 queries are significantly correlated in the USA and China. This concurs with the different timing of outbreaks of COVID-19 in the USA and China. In 2020, the search trends of 23 queries in China and in Wuhan are significantly correlated. This concurs with the fact that national attention has been highly focused on COVID-19 during the first five months of 2020. The analysis results indicate that the collective response to the COVID-19 pandemic exists in 2020.

**Figure S12:**
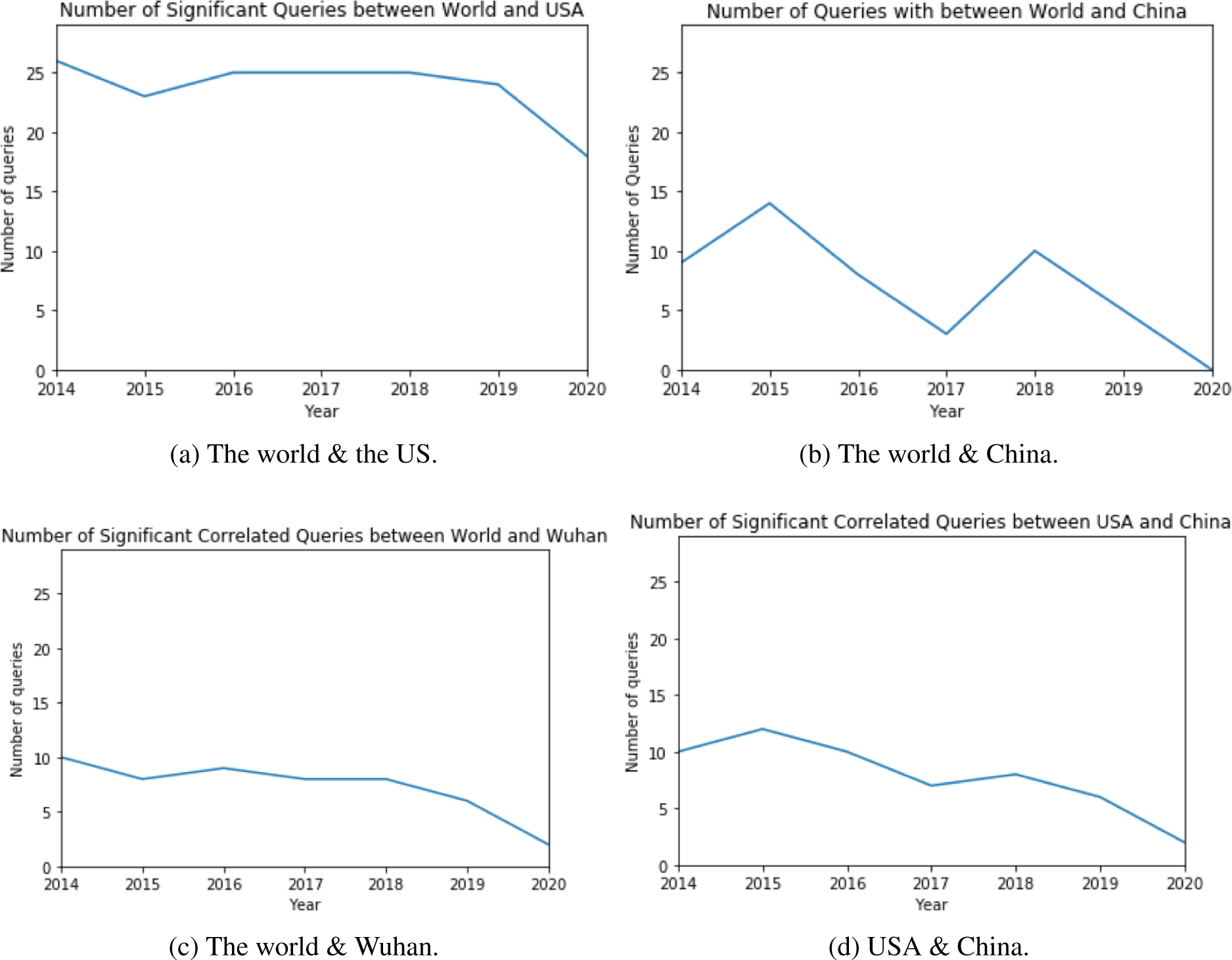
Number of significant queries by Spearman correlation.

### Search Data Analysis of Second Wave of Epidemic in Beijing

For over 30 search queries related to the COVID-19 pandemic, we analyze the daily search index trend in Beijing from March 1st, 2020 to July 20th, 2020 for each keyword, as Beijing was in the center of the epidemic. From the analysis, we are able to categorize them into two main categories, one group that has an abnormal rise in accordance with the pandemic shown in Figure S14, and another that has a relatively mild reaction shown in Figure S15. Some representative queries that belong to the first category include “nucleic acid,” “epidemic,” “mask,” “sore throat,” “fever,” “fatigue,” “pneumonia,” “diarrhea,” “coronavirus,” “Xinguan (stands for COVID),” “patient,” and “body temperature.” These words are mostly in high rankings in our previous correlation analysis, indicating consistency and effectiveness of analytical tools above. Let us take the word “epidemic” as an example. The searching trend of this word underwent stable declination throughout the period until the start of June, with the highest point between 1.5 and 2 at very first of March and the trough at around −1.5 on June 5th. This downward slope can be explained by a continuous period of zero confirmed cases in Beijing, and people gradually becoming less vigilant on the prevention of COVID-19. Yet, the search trend skyrockets significantly after June 10th, reaching its peak at more than 3.5 in merely 4 days. This nearly vertical line fits into the sudden onset of the second wave of COVID-19, as the first case was published in the news on June 11th, where the daily increase became abnormally high. This tremendous increase in search persisted for almost 14 days. The declination afterwards may be due to the fact that the number of newly confirmed cases fell as the government actively engaged in testing and containment measures in relevant locations in Beijing. The entire set of abnormal trends fits perfectly with the developing situation of the second wave of the pandemic, indicating the close-knit relationship between search trends and epidemic circumstances and the effectiveness of using search volume data as tools for exploring public reactions. Other search queries, such as “fever” and “nucleic acid,” show very similar patterns, with either parallel trend or decreasing trend over the past few months, and an obvious, exceptional, sudden rise starting at around June 10th or June 11th. One intriguing factor is that the keyword “diarrhea” is also in category one, with a significant high peak from June 10th to June 19th. However, “diarrhea” ranked relatively low in previous DCCA and DTW rankings, meaning that it’s not considered very pertinent to the virus by the general public. Yet, its search trend’s acute rise in the second wave of the epidemic differs from previous search trend changes in the first national wave. One reason may be that the epidemic center of Beijing is in Xinfadi, one of the most important food markets in that region, so people in Beijing may have become more concerned with food safety and food-related searches. Similar patterns of abnormal increase also applied to other common symptom words such as “fatigue” and “fever,” with their tremendous pump-ups in search volumes matching exactly with the period of the second wave of the pandemic in a range of approximately 15 days after the first announcement of the news, on June 11th, 2020.

**Figure S13:**
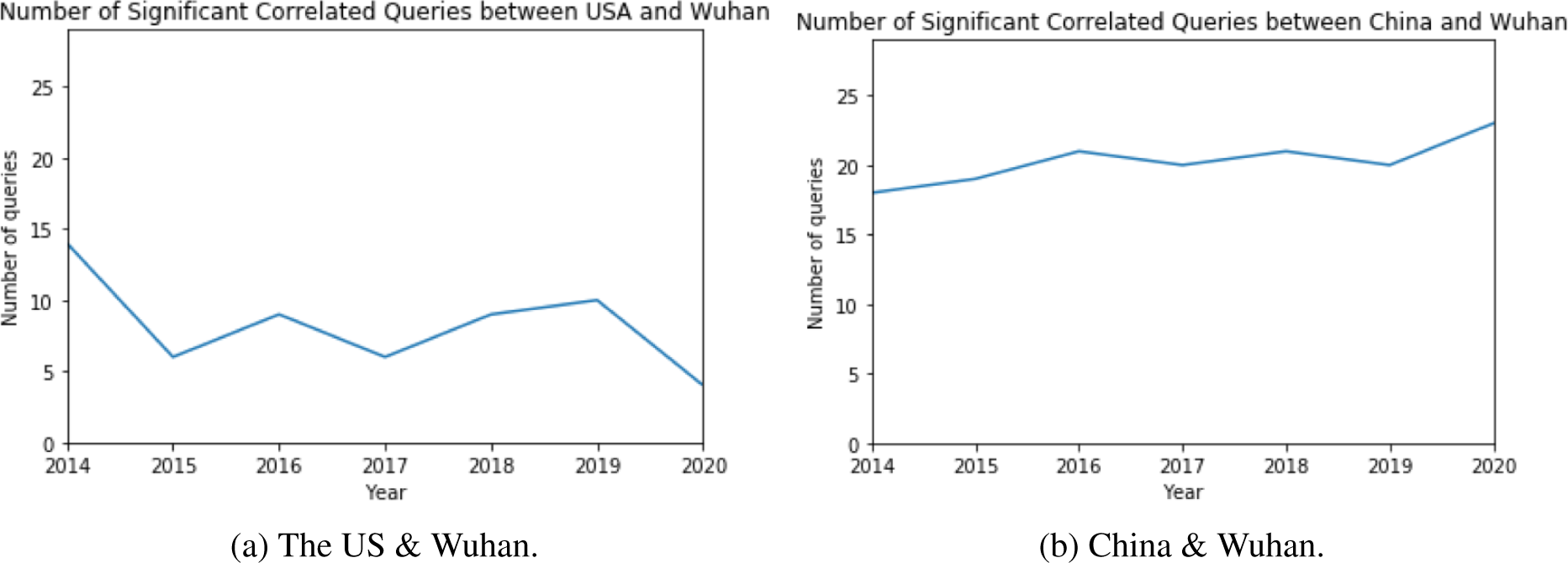
Number of significant queries by Spearman correlation.

**Figure S14:**
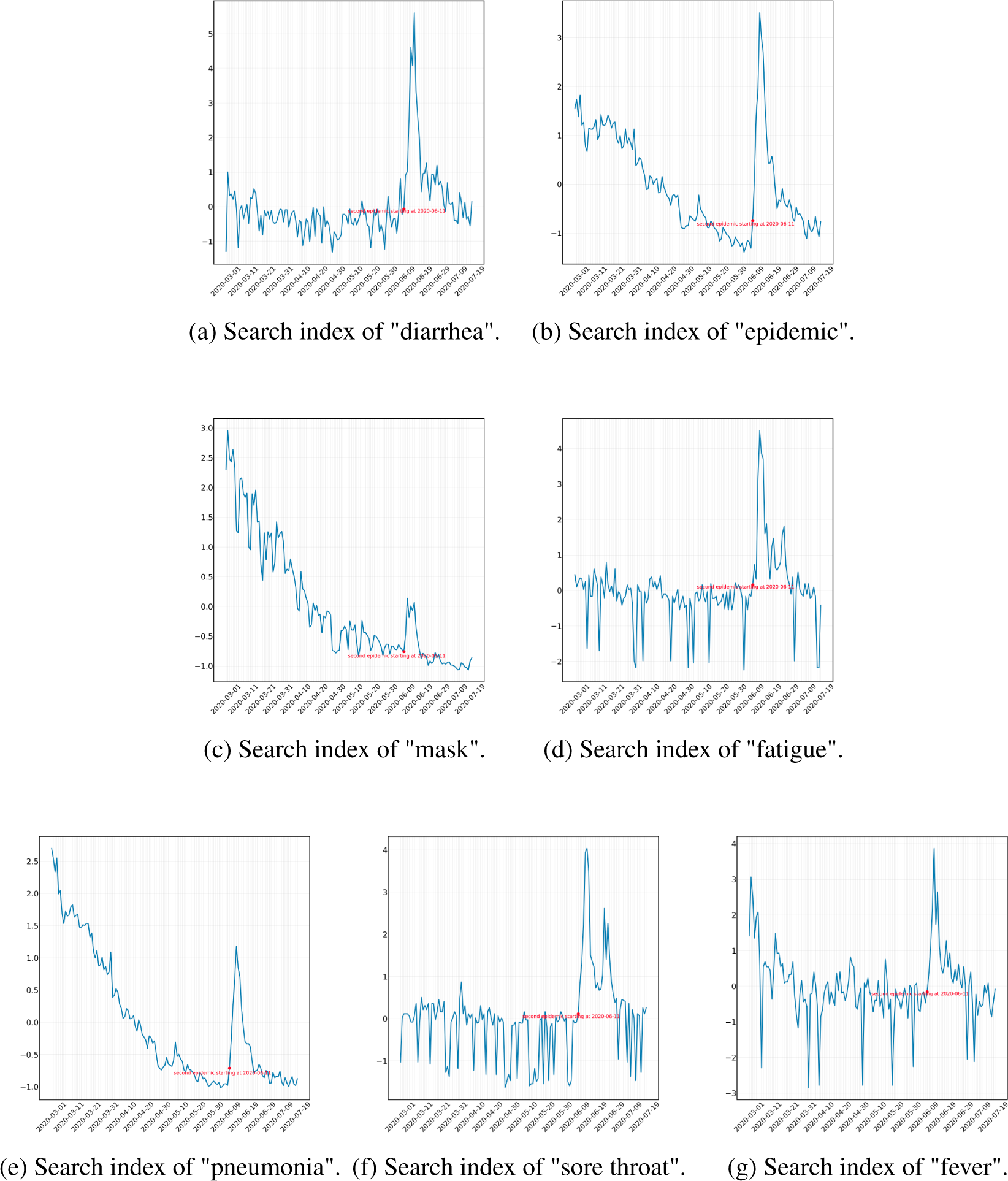
Analysis of search index data in Beijing from March 1st, 2020 to July 20th, 2020. The marked red points in the figures represent the starting date, i.e., June 11th, 2020, of the second COVID-19 pandemic in Beijing.

**Figure S15:**
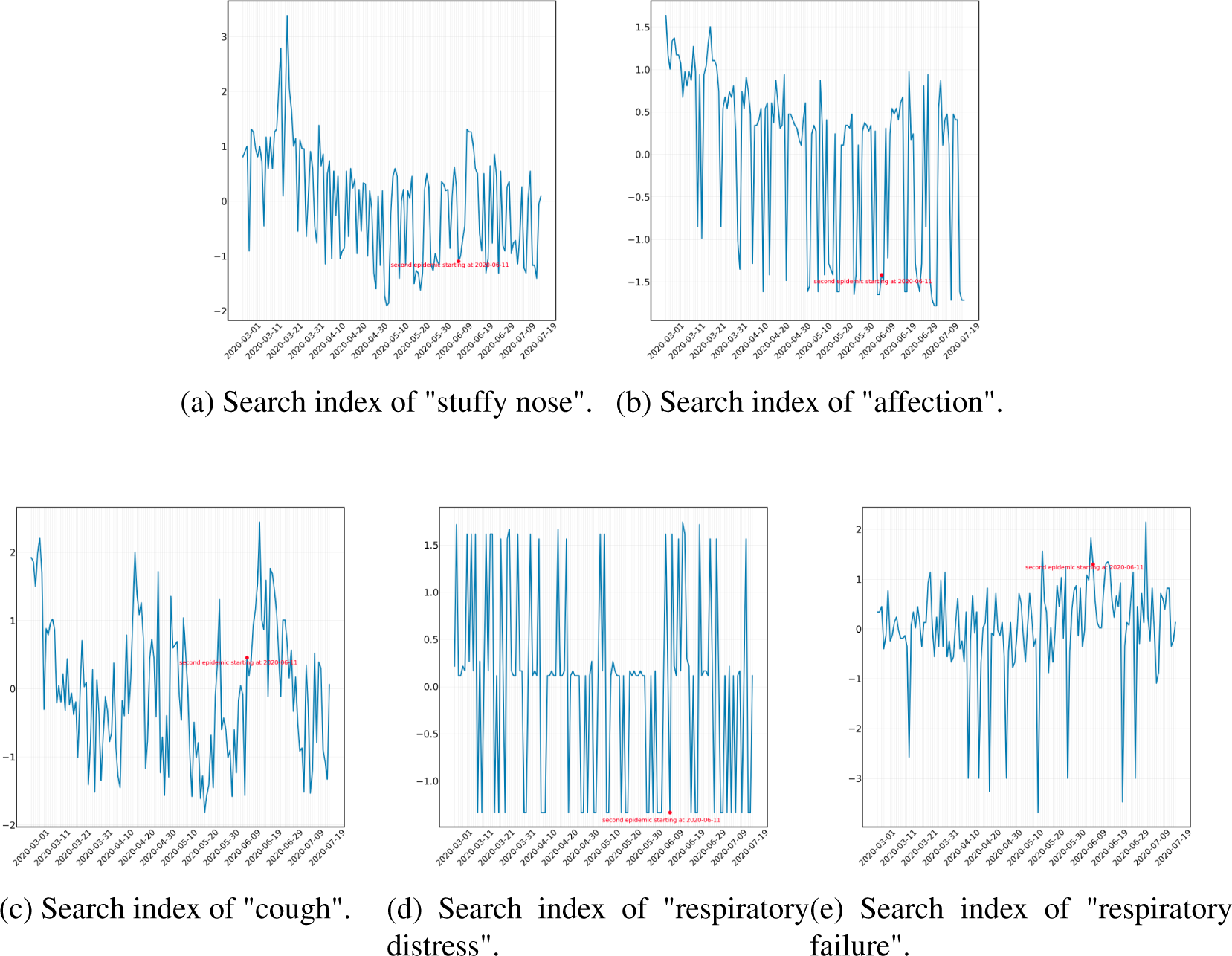
Analysis of search index data in Beijing from March 1st, 2020 to July 20th, 2020. The marked red points in the figures represent the starting date, i.e., June 11th, 2020, of the second COVID-19 pandemic in Beijing.

Furthermore, there are certain words that do not show abnormal patterns as discussed above, with no significantly abrupt increase in their overall trend from March to July 2020. For instance, the keyword “stuffy nose” has a searching trend with a peak in around the second half of March, following by a parallel trend throughout the period with normal daily fluctuations in a stable range. Although the peak at the week of June 11th, signaling the start of the second wave of the pandemic, was mildly wider than other fluctuations, it only showed a slightly higher search volume and a quick fall back to normal levels. It is hard to perceive significant traces of the second wave of the epidemic from the diagram directly. In similar fashion, search queries such as “respiratory failure” and “respiratory distress” display no significant deviation from the general trend since March, with no especially peculiar shape of fluctuations. Although these words are considered relevant to COVID-19, these symptoms are more related to severely ill patients. Consequently, it is reasonable to see no significant change in these keywords, since Beijing has had only a few critically ill patients, compared to the first national wave of the pandemic, resulting in many fewer people searching for these symptoms of serious illness. Other keywords, including “respiratory symptom,” “dehydration,” and “affection” also show no obvious change in search volume trend during this second epidemic in Beijing. One crucial search query, “cough,” also displays less significant change in trend compared to other keywords. Upon closer examination, there is a hump from early June to the end of June, with the highest peak on June 11th. Yet most peaks around the period from June 11th to July 20th are still at equivalent levels, compared to the high points in April and May, and thus the second wave of Beijing epidemic outbreak did not cause a tremendous increase in search volume for “cough.” Among keywords, “cough” is also abnormal. It varies from other search trend changes. In the first national wave of the epidemic, its ranking is very high by DCCA and DTW rankings; but in the second wave, the public did not show a strong reaction by searching significantly more frequently for “cough.” Comprehensive reasons are still under investigation, but it may be due to the fact that the confirmed case number is not very high, due to the efficient measures to contain the spread. Another possibility to consider is that public education had already covered early on that coughs could be a symptom, while the knowledge that other symptoms, such as diarrhea, could derive from the new novel coronavirus emerged later. In fact, our techniques lend credence to the notion that looking proactively at what diagnostic terms are being searched for could potentially help identify symptoms that are not already known to be part of the pandemic. Notwithstanding this word, most other words show strong consistency with our previous analysis, as search queries with ranking higher than the 15^*th*^ place are mostly in category one.

### Population in the Hospitals of Wuhan

In this section, we present the figures of population data in shelter (fangcang) hospitals (*39*) and Huanan seafood market. We calculate the total population of shelter hospitals and the population of Huanan seafood market. As shown in Figure S16, the figure of shelter hospitals exhibits highly similar patterns with the figure of hospitals described in the Section, Analysis of Population Densities in Hospitals of Wuhan. After areas were used to build shelter hospitals, steps to close off and manage ingress and egress were implemented, meaning that strict restrictions on people entering and leaving were enacted; thus, the population data fell off enormously. There is no significantly aberrant deviation of trend in population data of late 2019 in reference to data of late 2018. This conclusion also applies to patterns of population densities in Huanan seafood market as shown in Figure S17, as there is no abnormal deviation from paths in the previous year. Consequently, the latter part of 2019 did not witness any aberrant signal that could link to the collective response to the COVID-19 pandemic.

**Figure S16:**
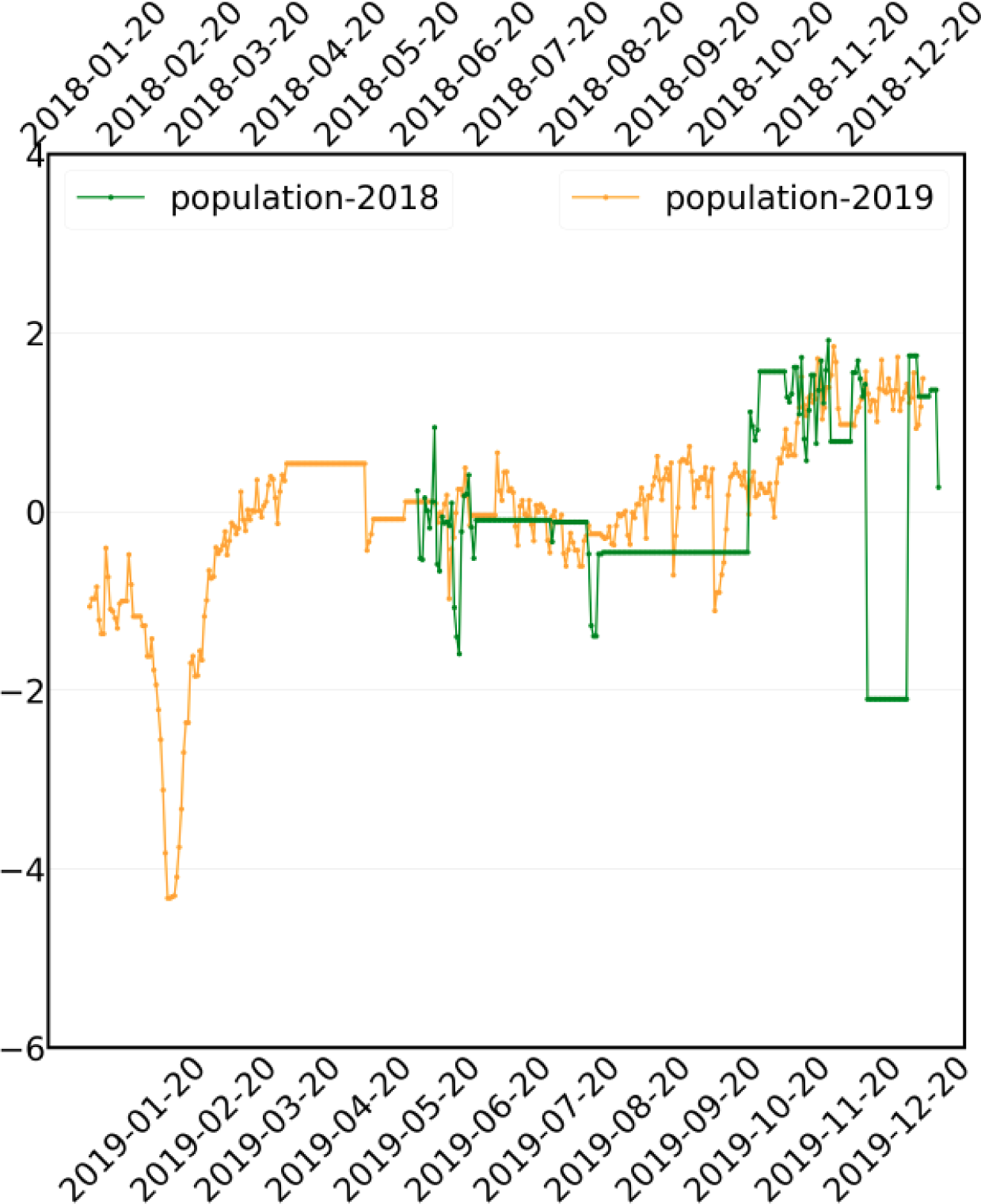
Population data of shelter hospitals in Wuhan.

**Table S1:** Keyword ranking in Wuhan. “NCP” represents “Novel Coronavirus Pandemic.”

| Rankings | Keywords (based on DTW) | Keywords (based on DCCA) |
| --- | --- | --- |
| 1 | COVID | COVID |
| 2 | Epidemic situation | Epidemic situation |
| 3 | Nucleic acid | Nucleic acid |
| 4 | Mask | NCP |
| 5 | Pneumonia | Hypoxemia |
| 6 | Sterilization | Sterilization |
| 7 | Asthenia | Infection |
| 8 | Sore throat | Contagion |
| 9 | Infection | Pneumonia |
| 10 | NCOV | Inflammation |
| 11 | Medical case | Medical case |
| 12 | Tachypnea | Runny nose |
| 13 | Hypoxemia | Asthenia |
| 14 | Respiratory distress | Sore throat |
| 15 | Body temperature | Body temperature |
| 16 | Fever | Respiratory distress |
| 17 | NCP | Mask |
| 18 | Inflammation | Dehydration |
| 19 | Diarrhea | Dyspnea |
| 20 | Contagion | Tachypnea |
| 21 | Dry cough | Diarrhea |
| 22 | SARS | NCOV |
| 23 | Dyspnea | Respiratory failure |
| 24 | Dehydration | Fever |
| 25 | Runny nose | Dry cough |
| 26 | Respiratory failure | Cough |
| 27 | Stuffy nose | Stuffy nose |
| 28 | Cough | SARS |

**Table S2:**
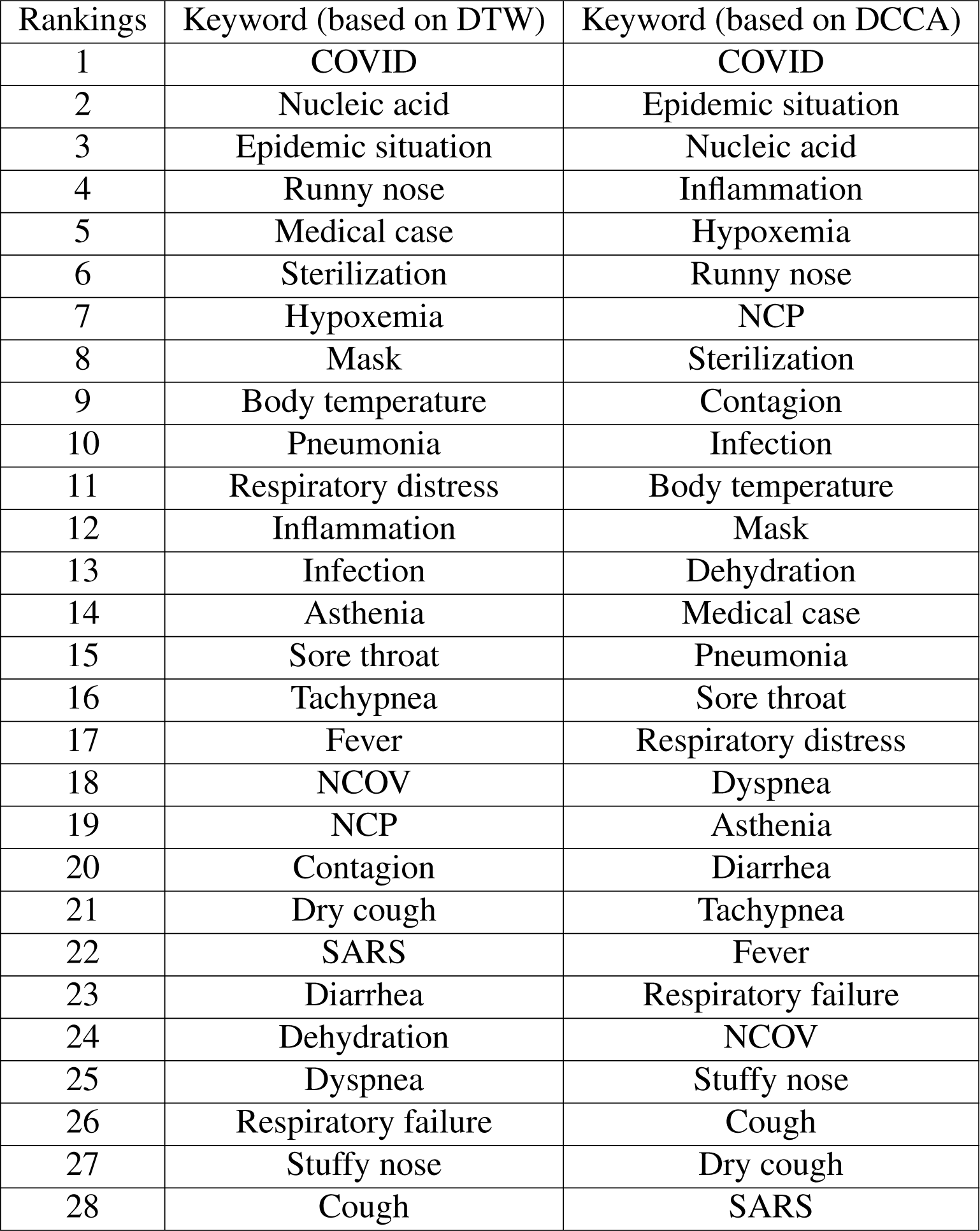
Keyword ranking in China. “NCP” represents “Novel Coronavirus Pandemic.”

**Table S3:** Keyword ranking in US. “NCP” represents “Novel Coronavirus Pandemic.”

| Rankings | Keyword (based on DTW) | Keyword (based on DCCA) |
| --- | --- | --- |
| 1 | COVID | Mask |
| 2 | Mask | COVID |
| 3 | Sterilization | Sterilization |
| 4 | Body temperature | Body temperature |
| 5 | SARS | Food refusal |
| 6 | NCOV | Respiratory failure |
| 7 | Epidemic situation | Respiratory distress |
| 8 | Food refusal | Hypoxemia |
| 9 | Dry cough | Epidemic situation |
| 10 | Fever | Nucleic acid |
| 11 | Respiratory distress | Asthenia |
| 12 | Asthenia | Dry cough |
| 13 | Inflammation | Dehydration |
| 14 | Dyspnea | Dyspnea |
| 15 | Sore throat | Hypoxemia |
| 16 | Diarrhea | SARS |
| 17 | Runny nose | Medical case |
| 18 | Hypoxemia | Diarrhea |
| 19 | Pneumonia | NCP |
| 20 | Cough | Inflammation |
| 21 | Contagion | Fever |
| 22 | Dehydration | Sore throat |
| 23 | Stuffy nose | Stuffy nose |
| 24 | Asthenia | Runny nose |
| 25 | Tachypnea | Pneumonia |
| 26 | Nucleic acid | Cough |
| 27 | Medical case | NCOV |
| 28 | NCP | Contagion |

**Table S4:** Keyword ranking all over the world. “NCP” represents “Novel Coronavirus Pandemic.”

| Rankings | Keyword (based on DTW) | Keyword (based on DCCA) |
| --- | --- | --- |
| 1 | COVID | COVID |
| 2 | Mask | Mask |
| 3 | Sterilization | Sterilization |
| 4 | Body temperature | Body temperature |
| 5 | SARS | Food refusal |
| 6 | Food refusal | Respiratory distress |
| 7 | Epidemic situation | Respiratory failure |
| 8 | NCOV | Hypoxemia |
| 9 | Respiratory distress | Epidemic situation |
| 10 | Respiratory failure | Dyspnea |
| 11 | Dyspnea | Dry cough |
| 12 | Inflammation | Medical case |
| 13 | Contagion | Dehydration |
| 14 | Sore throat | Nucleic acid |
| 15 | Fever | Tachypnea |
| 16 | Pneumonia | Sore throat |
| 17 | Runny nose | Inflammation |
| 18 | Hypoxemia | Asthenia |
| 19 | Sore throat | Fever |
| 20 | Diarrhea | Runny nose |
| 21 | Tachypnea | SARS |
| 22 | Asthenia | Diarrhea |
| 23 | Medical case | Stuffy nose |
| 24 | Cough | Contagion |
| 25 | Stuffy nose | NCP |
| 26 | Nucleic acid | Pneumonia |
| 27 | NCP | Cough |
| 28 | Asthenia | NCOV |

**Figure S17:**
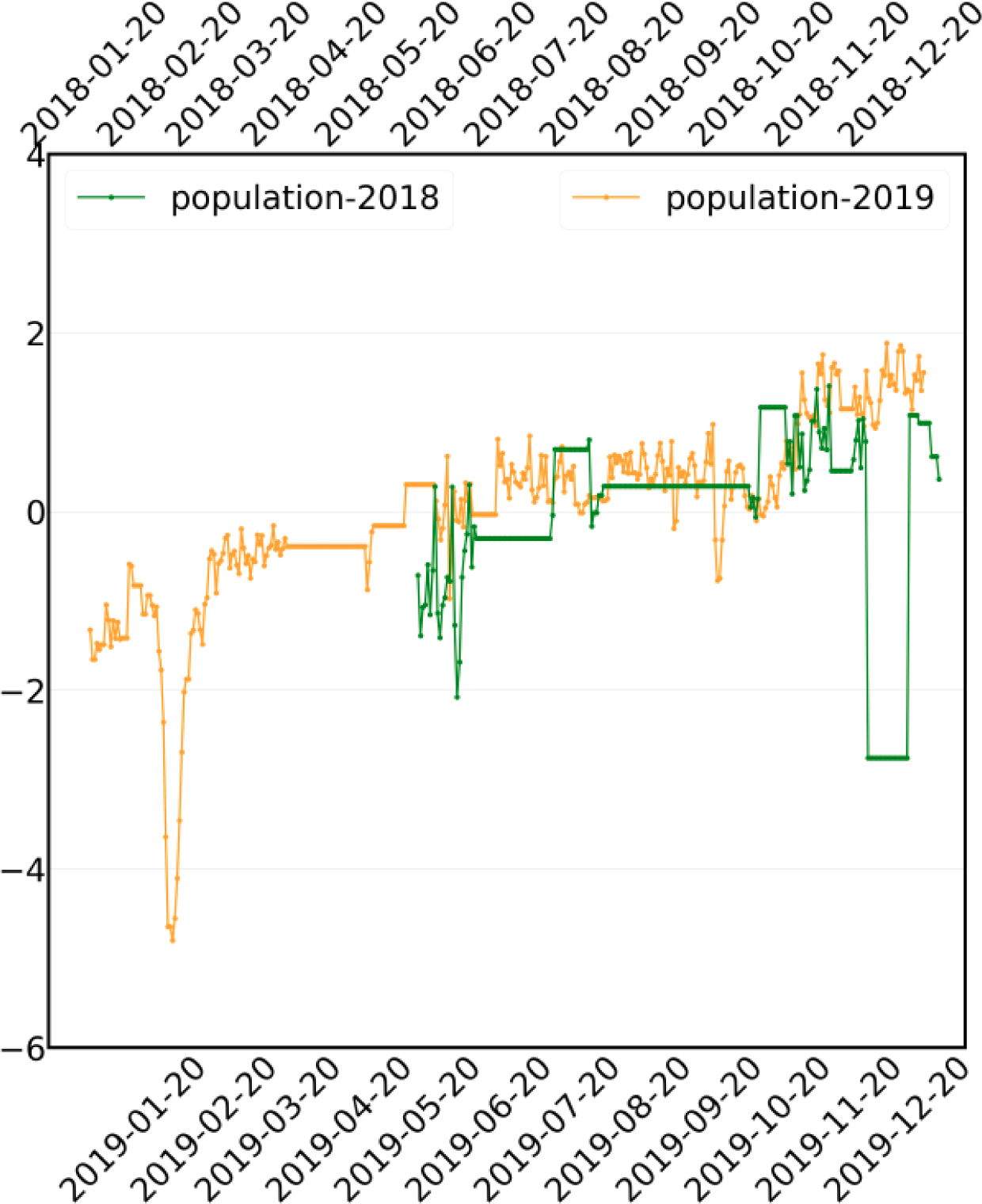
Population data of seafood markets in Wuhan.

